# Which surface treatment improves the long-term repair bond strength of aged methacrylate-based composite resin restorations? A systematic review and network meta-analysis

**DOI:** 10.1101/2022.06.07.22276106

**Authors:** Mahdi Hadilou, Amirmohammad Dolatabadi, Morteza Ghojazadeh, Hossein Hosseinifard, Parnian Alizadeh Oskuee, Fatemeh Pournaghi Azar

**Author notes:** **Correspondence to:** Dr. Fatemeh Pournaghi Azar, DDS, MSD, Golgasht Blvd, Faculty of Dentistry, Tabriz University of Medical Sciences, Tabriz, Iran, Postal code: 51666-16471.

## Abstract

This study aimed to investigate the effect of common surface treatments on the long-term repair bond strength of the aged methacrylate-based composite resin restorations. Also, provide their rankings and two-by-two comparison. In-vitro studies evaluating the methacrylate-based composite resins subjected to rigorous aging procedures before and after being repaired with a new composite were included. A frequentist network meta-analysis was carried out using a random-effects model. P-scores were used to rank the efficacy of the surface treatments. Also, the global and node-split inconsistencies were evaluated. Web of Science, PubMed/Medline, Scopus, and Embase databases were searched until November 11, 2021. Application of diamond bur + silane + total-etch adhesive (shear MD 41.12 MPa, 95% CI 29.02 to 53.21, P-score 0.99; tensile MD 29.36 MPa, 95% CI 12.36 to 46.35; P-score 0.69), and air abrasion with silica-coated alumina + silane + total-etch adhesive (shear MD 16.29 MPa, 95% CI 6.54 to 26.05, P-score 0.66; tensile MD 33.86 MPa, 95% CI 16.17 to 51.54; P-score 0.82) produced the highest (micro)tensile and (micro)shear bond strengths compared to abrasive paper, according to two network meta-analyses containing 22 investigations. There has been no variation comparing self- and total-etch adhesives. Further, mechanical surface treatments should be used alongside the chemical adhesive agents to yield stronger bonds. It is possible to achieve acceptable repair bond strength using common dental clinic equipment. Therefore, clinicians could consider repairing old composite resins rather than replacing them.

## Introduction

Composite resin restorations must be repaired or replaced, over different time periods following placement due to secondary caries, cracks, marginal discoloration, gaps, cosmetic improvements, shape corrections, and proximal contact closures [1-3].

Based on the extent of the damage and the seriousness of the defect, repair may be a viable option to replacement. Recent advances in adhesive dentistry [4], and equivalent or even superior clinical survival rates of repaired composite resin restorations than replaced ones [5-7] have led to a paradigm shift from full replacement of slightly defective old composite resin restorations towards repairing the defected portion in accordance with minimally invasive dentistry [6, 8].

This approach offers a number of benefits, including preservation of sound tooth structure, prevention of iatrogenic manipulation of pulp tissue [9], time savings, and decreased clinical treatment expenses for individuals [10]. It also is more accepted across patients than replacement [11, 12]. However, due to the Gradual deterioration of the structure of composite resin restorations following water absorption in the oral environment [13, 14], and a decrease in chemical surface activity owing to the unavailability of an oxygen-inhibited layer that provides the unreacted residual monomers as a key element in the bonding procedure [15, 16], there are obstacles to attain a satisfactory bond between new and old composite resin restorations.

Patient-related variables (oral hygiene, caries risk, and occlusal forces) and dentist-related ones (experience, technique, and material selection) impact the clinical performance of fresh to old composite resin bond strength [17-19]. The surface treatment protocols performed on old composite resins also play a significant role in determining the durability of the repair bond. In-vitro research findings had also utilized a variety of surface treatment methods, such as roughening with abrasive papers [20-23], diamond burs [24-26], Al_2_O_3_ particles [3, 22, 27], silica coated particles [28-30], and lasers [31, 32] as mechanical; and numerous bonding mechanisms, adhesives, and salinization as chemical surface treatments [3, 22, 23, 27], solely or in combination to overcome the bonding obstacles. The main objective of such surface modifications is to produce a strong bond through the micromechanical interlocking between fresh and old composite resins.

The quantity and type of enforced aging also influences the durability and sustainability of repair bond strength of aged composite resins. This process of aging should reflect the fundamental mechanical and chemical properties of the oral environment, such as temperature and pH variations, masticatory forces, and oral habits [33]. Valente et al. presented in 2016 the most recent systematic review and meta-analysis investigating the influence of various surface treatments on the repair bond strength of aged composite resins. Their results were largely derived on relatively short-term static water storage aging tests; thus, they would not have adequately described the long-term repair bond characteristics [34].

To the best of our knowledge, a systematic review and network meta-analysis assessing the influence of common surface treatment approaches on the long-term repair bond strength of properly aged composites resin restorations has still not been conducted. Further, in the lack of head-to-head trials, no research has compared the common surface treatments.

A network meta-analysis offers clinicians with such a rating of surface treatments that may motivate them to repair composite resin restorations with minor defects rather replacing them. In addition, a well-established guideline has still not been developed. This systematic review and network meta-analysis was conducted to offer an explanation for, “Which surface treatment strategy improves the long-term repair bond strength of aged methacrylate-based composite resin restorations?”

## Materials and methods

This research is conducted in accordance with the Cochrane Handbook for Intervention Reviews [35] and Preferred Reporting Items for Systematic Reviews and Meta-Analyses (PRISMA) extension for network meta-analyses [36] (Supplementary material 1). The study protocol is registered in the International Prospective Register of Systematic Reviews database (Registration code CRD42022308586).

### Eligibility Criteria

The studies were included based on the following PICO:

- *Population:* Appropriately aged direct or indirect methacrylate-based composite resins (thermocycling at least for 5000 cycles or stored in water at least for 4 weeks). The aging procedure should have been caried out both before surface treatment (primary aging) and after repairing the aged composite resin (secondary aging). The control group’s composite resins must have likewise undergone the aging process.
- *Intervention:* Chemical and physical surface treatments including roughening with lasers, diamond burs, Al_2_O_3_ or silica-coated alumina particles, administration of adhesives or silanes, etc. The included papers required a minimum of two comparison groups.
- *Comparison:* In most published investigations, grinding using abrasive paper was deemed the control group since it is a part of the standard sample preparation procedure.
- *Outcome:* Repair bond strengths including tensile, shear, micro tensile, and micro shear bond strengths. Studies using non-methacrylate-based composite resins such as resin nanoceramics, polymer-infiltrated ceramics and silorane-based composite resins, or uncommon aging methods such as UV aging were excluded. Only English-language publications were included.

### Information Sources and Search

MH conducted a thorough search up to November 11, 2021, in 4 databases, namely PubMed/Medline, Embase, Scopus, and Web of Science, using the search strategies outlined in Supplementary material 2. The search strategies consisted of free keywords, Medical Subject Headings (MeSH) terms, and Emtree keywords, combined with the OR and AND Boolean operators. The asterisk (*) was used to increase the searching accuracy. There was a search of ProQuest Dissertation & Theses for gray literature. Additionally, the references of the included studies were searched for related literature.

### Study Selection and Data Extraction

Two researchers (MH and AD) reviewed the papers separately based on eligibility criteria. First on the basis of the titles and abstracts, next the full-texts. When conflicts arose, a third researcher (PAO) was consulted. The extraction table was created during the pilot stage of the study including 10 articles. The extraction table included the author, year, type of aged and repaired composites, types of primary and secondary aging, total sample for each group, the type of surface treatment (chemical and mechanical), the method of bond strength test and the related outcomes.

Two researchers (MH and AD) executed the data extraction in duplicate. In the instance of missing information, the study authors were contacted; if no response was obtained, the data were retrieved conservatively. For example, if a spectrum was given as outcome of each study group, the bottom end of the spectrum was used to establish the group’s sample size. If insufficient information were provided, the research would be discarded.

If several doses or levels of a surface treatment were employed in a paper, the only one most often used in the clinical environment was included in the extraction table. In cases where the bond strength of various kinds of composite resins was recorded, the information of all of them was also included in the extraction table. If a study assessed the bond strength of a composite resin using a single surface treatment but under multiple aging methods, the strongest aging technique was reported in the extraction table.

### Risk of bias assessment

The risk of bias assessment was adapted from The Cochrane Collaboration’s tool for assessing risk of bias [37] and previously published studies [34, 38]. Two researchers (MH and AD) conducted the risk of bias assessment separately, resolving discrepancies by consulting the third researcher (FPA). The risk of bias graph and summary was created using the robvis visualization instrument [39].

### Statistical Analyses

MG and HH conducted a frequentist network meta-analysis employing a random-effects model utilizing R software’s net meta module (version 3.6.2; R Foundation for Statistical Computing). Every node indicated an intervention from the articles included in the analysis. The lines linking the nodes reflected present head-to-head comparisons, and their thickness was proportional to the number of studies that examined these comparisons. The surface treatments were ranked according to their respective P-scores. Higher P-scores demonstrate a higher likelihood that the intervention is superior to other comparisons [40]. The network-level inconsistency (global inconsistency) was presented by I^2^ [41]. In addition, node-split inconsistency was examined by comparing the direct and indirect evaluations. If there was a significant discrepancy (P<0.05) in the direct and indirect evaluations, the direct evaluation was reported as the pooled estimate in the league tables.

To identify any publication bias or small study effect, comparison-adjusted funnel plots were created for all possible comparisons with abrasion paper serving as the control group.

In the case of reporting the bond strength of multiple types of methacrylate-based composites with the identical intervention, the mean ± standard deviation of these groups was merged into one group. The adhesives were classified as either self-etch or total-etch. In the case of using multiple mechanical surface treatments on a single group of composite resins, all treatments were reported in the extraction table. However, only the dominant physical surface treatment depicted that group over the abrasive paper in the network meta-analysis. Further, acid etching was not regarded a surface treatment since it is primarily employed for surface cleaning [42].

## Results

The search yielded 5062 papers, of which 2225 were duplicates. After screening based on titles and abstracts, 175 papers were chosen for full-text examination, of which 22 fulfilled the inclusion criteria and were included in the network meta-analysis. The excluded articles and their justifications may be found in Supplementary material 3. The PRISMA flowchart is shown in Figure 1.

**Figure 1:**
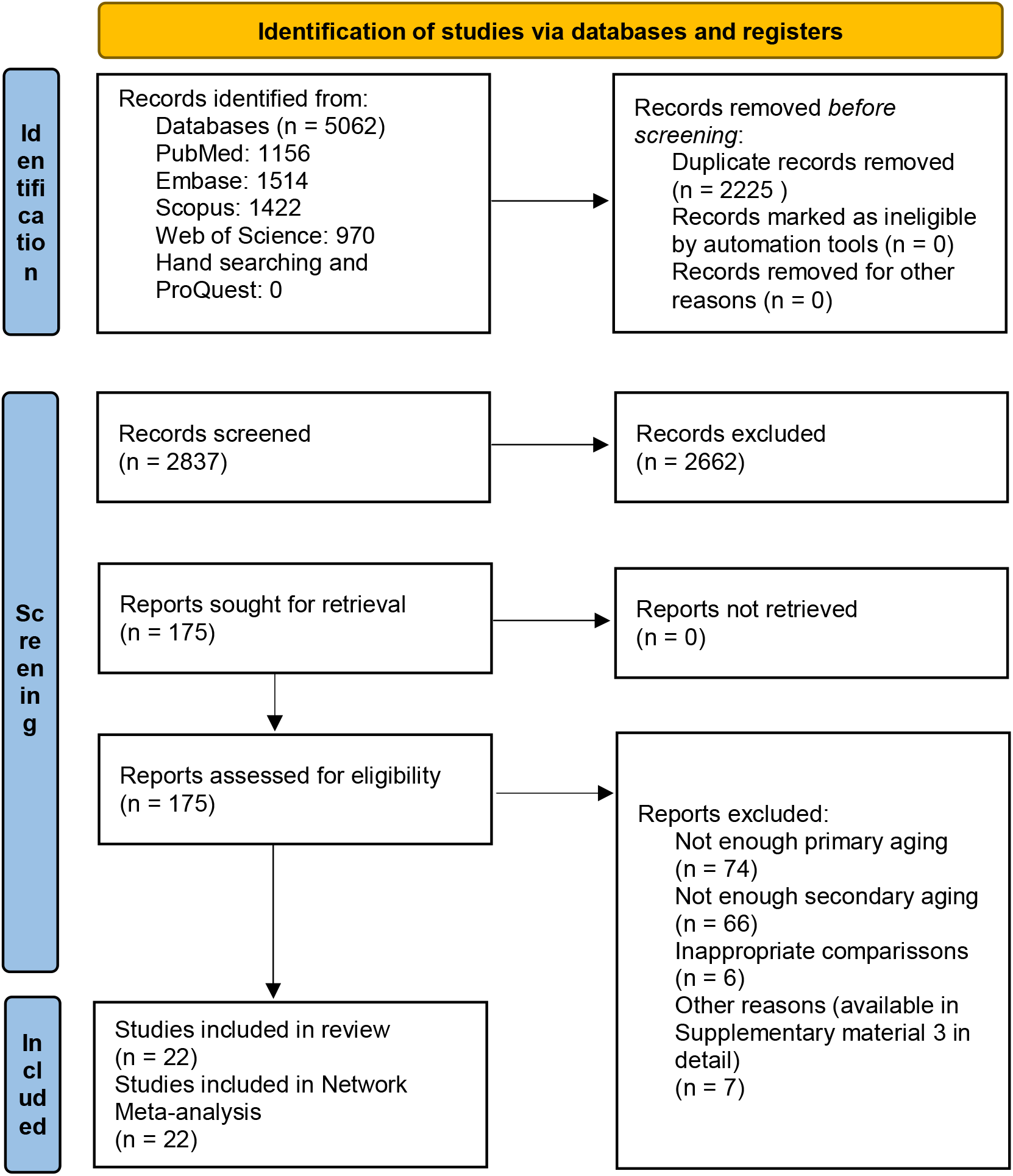
PRISMA flowchart.

### Characteristics of included studies

All twenty-two included studies had in-vitro lab settings. The publishing dates of the included papers spanned from 2007 to 2021. The sample sizes varied between 5 [43] to 61[30] in each study. The aging methods included liquid storage (water [3, 9, 27, 44, 45] and saliva [46]), thermocycling [20, 22-25, 29, 31, 32, 42, 47], or a combination of the two [21, 26, 28, 30, 43, 48]. Seven studies [22-25, 30, 43, 46] provided micro-tensile bond strength, four [3, 9, 44, 47] indicated micro-shear bond strength, ten [20, 21, 26-29, 31, 32, 42, 48] gave shear bond strength, and only one [45] evaluated tensile bond strength. A full extraction table may be found in Supplementary material 4.

### Risk of bias assessment

All of the studies had different levels of bias (Supplementary material 5). The absence of blinding the testing machine operator was the most biased field of the included research. In addition, sample size calculation seemed to be mostly arbitrary across the studies. Furthermore, standard preparation of specimens (by single operator), and standard specimen selection (the procedure of identifying and excluding defective specimens) domains posed a significant risk of bias (Figure 2).

**Figure 2:**
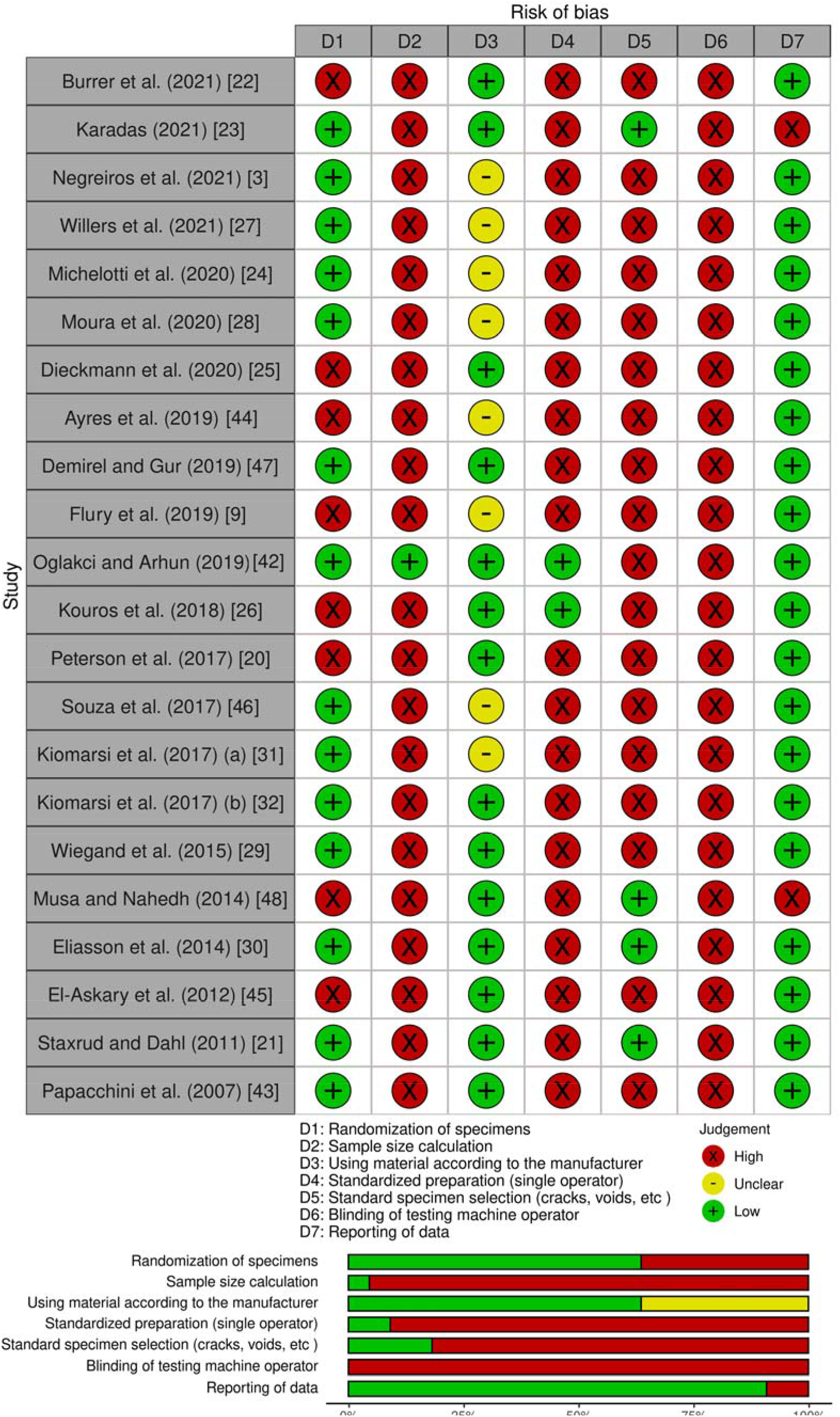
Risk of bias summary.

### Network meta-analysis results

Twenty-two studies were included in the network meta-analysis. The global network heterogeneity was moderate for (micro)tensile networks (I^2^ = 56.9) and substantial for (micro)shear network (I^2^ = 94.1). The following surface treatments were used alone or in combination: 1) AP: abrasive paper 2) No: no surface treatment 3) SB: sandblasting with Al_2_O_3_ particles 4) SI: silane 5) total: total-etch adhesive 6) self: self-etch adhesive 7) AR: argon plasma 8) DB: diamond bur (with coarse particles for surface treatment purpose) 9) SC: air abrasion with silica-coated Al_2_O_3_ particles 10) FRC: flowable resin composite 11) LA: laser. Totally, 24 surface treatments were included in the (micro)shear network (Figure 3) and 24 in the (micro)tensile network (Figure 4). Supplementary material 6 contains the league tables.

**Figure 3:**
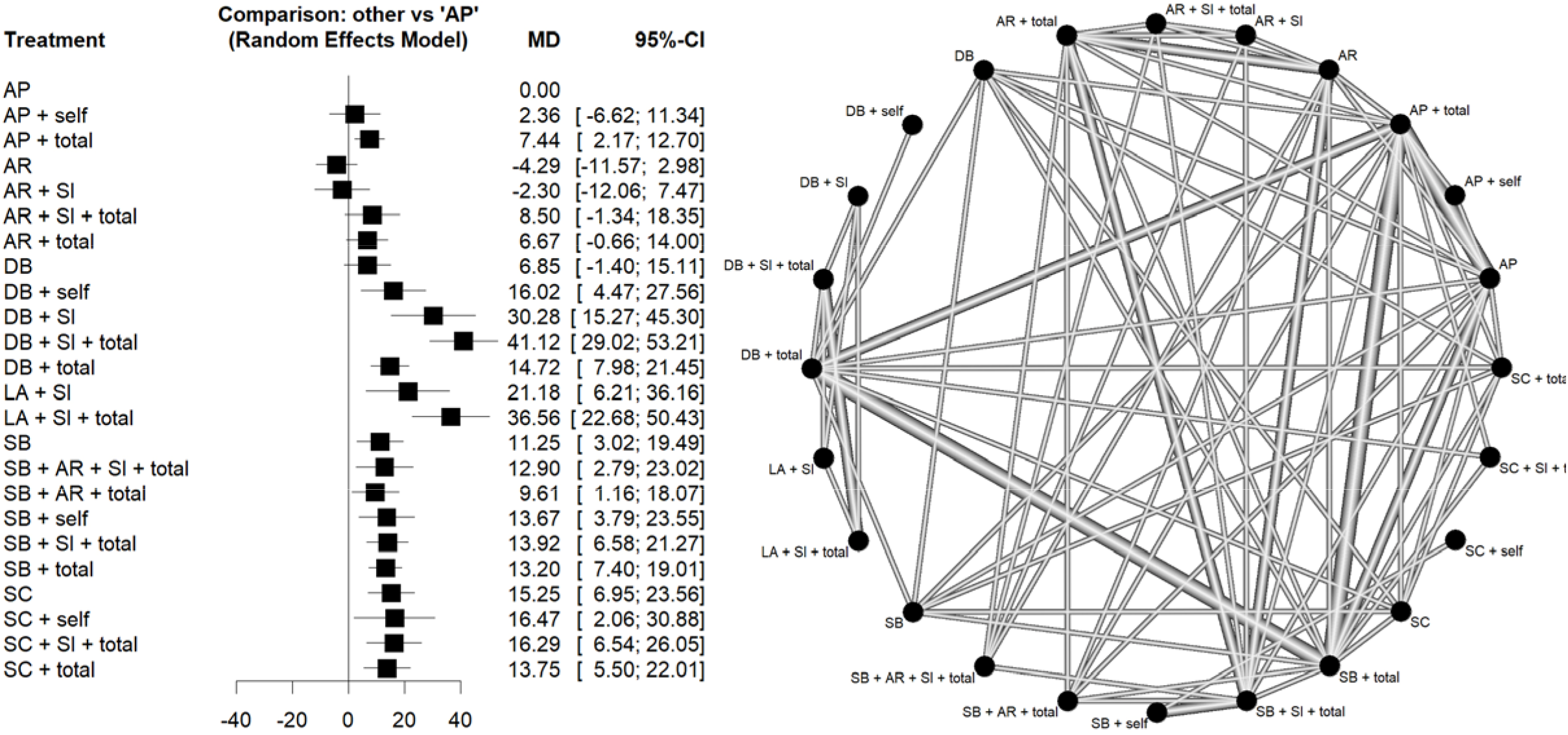
Network map of surface treatments introduced into the network meta-analysis for (micro)shear bond strength by random-effects model. (AP: abrasive paper; No: no surface treatment; SB: sandblasting with Al_2_O_3_ particles; SI: silane; total: total-etch adhesive; self: self-etch adhesive; AR: argon plasma; DB: diamond bur; SC: air abrasion with silica-coated Al_2_O_3_ particles; FRC: flowable resin composite; LA: laser)

**Figure 4:**
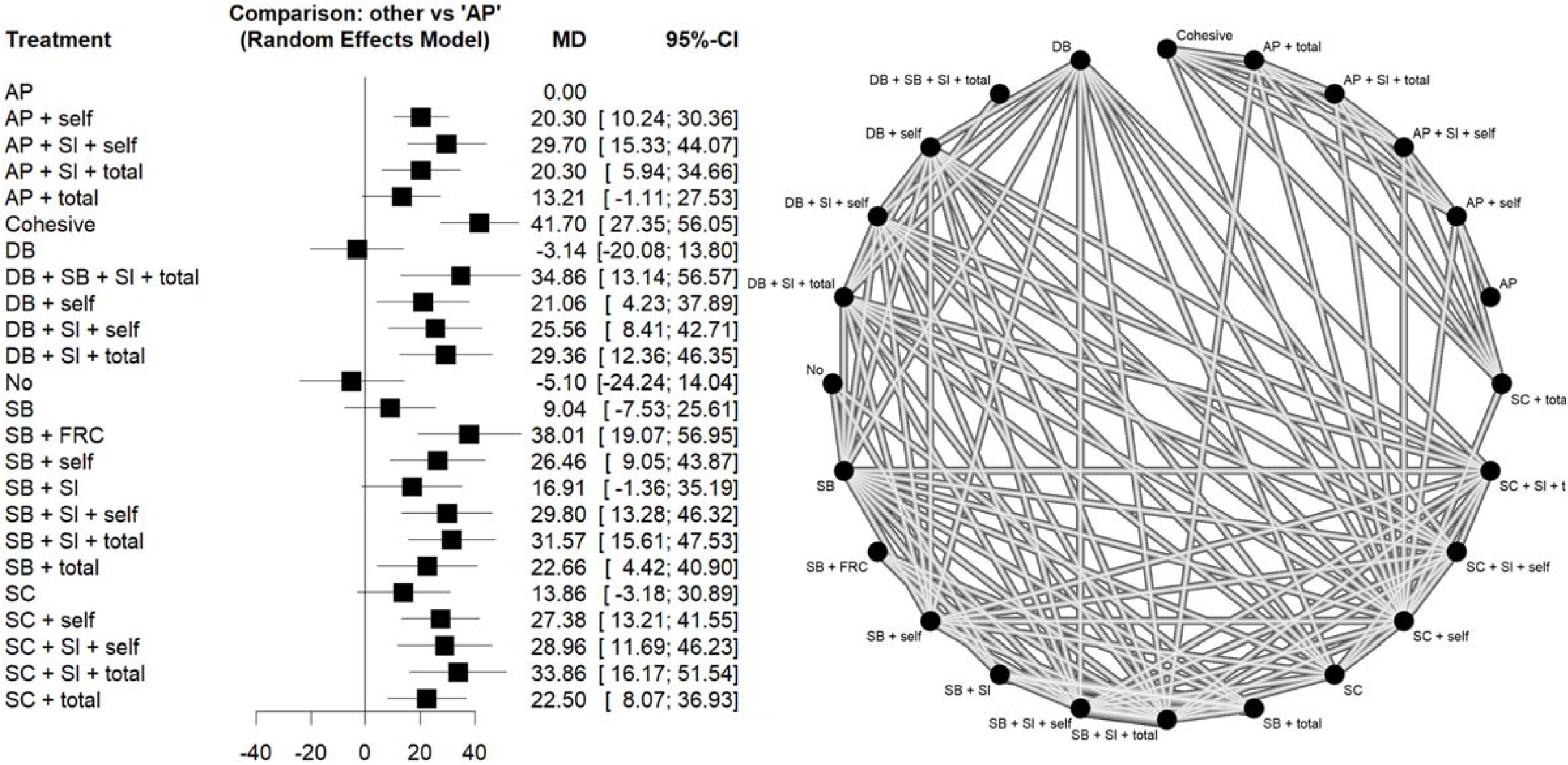
Network map of surface treatments introduced into the network meta-analysis for (micro)tensile bond strength by random-effects model. (AP: abrasive paper; No: no surface treatment; SB: sandblasting with Al_2_O_3_ particles; SI: silane; total: total-etch adhesive; self: self-etch adhesive; AR: argon plasma; DB: diamond bur; SC: air abrasion with silica-coated Al_2_O_3_ particles; FRC: flowable resin composite; LA: laser)

The forest plots are shown in Figures 3 and 4, and Table 1 contains the associated P-scores. Most head-to-head comparisons were based on a single study. The surface treatment with the greatest chance of producing higher (micro)shear bond strength was DB+SI+total (P-score = 0.99), preceded by LA+SI+total (P-score = 0.95), and DB+SI (P-score = 0.90).

**Table 1:** Surface treatment ratings by corresponding P-scores.

| Rank | Tensile |  | Shear |  |
| --- | --- | --- | --- | --- |
|  | Treatment | P-score | Treatment | P-score |
| 1 | Cohesive | 0.9595 | DB+SI+total | 0.9953 |
| 2 | SB+FRC | 0.9058 | LA+SI+total | 0.9552 |
| 3 | SC+SI+total | 0.8279 | DB+SI | 0.9002 |
| 4 | DB+SB+SI+total | 0.8176 | LA+SI | 0.7602 |
| 5 | SB+SI+total | 0.7740 | SC+SI+total | 0.6698 |
| 6 | SB+SI+self | 0.7114 | SC+self | 0.6509 |
| 7 | AP+SI+self | 0.7066 | DB+self | 0.6465 |
| 8 | DB+SI+total | 0.6907 | SC | 0.6382 |
| 9 | SB+SI+self | 0.6775 | DB+total | 0.6213 |
| 10 | SC+self | 0.6266 | SB+SI+total | 0.5897 |
| 11 | SB+self | 0.5875 | SC+total | 0.5730 |
| 12 | DB+SI+self | 0.5546 | SB+self | 0.5658 |
| 13 | SB+total | 0.4642 | SB+total | 0.5515 |
| 14 | SC+total | 0.4623 | SB+AR+SI+total | 0.5372 |
| 15 | DB+self | 0.4036 | SB | 0.4583 |
| 16 | AP+self | 0.3922 | SB+AR+total | 0.3891 |
| 17 | AP+SI+total | 0.3919 | AR+SI+total | 0.3464 |
| 18 | SB+SI | 0.3028 | AP+total | 0.2831 |
| 19 | SC | 0.2320 | DB | 0.2765 |
| 20 | AP+total | 0.2158 | AR+total | 0.2615 |
| 21 | SB | 0.1507 | AP+self | 0.1547 |
| 22 | AP | 0.0710 | AP | 0.0887 |
| 23 | DB | 0.0427 | AR+SI | 0.0616 |
| 24 | No | 0.0311 | AR | 0.0253 |

AP: abrasive paper; No: no surface treatment; SB: sandblasting with $\text{Al}_2\text{O}_3$ particles; SI: silane; total: total-etch adhesive; self: self-etch adhesive; AR: argon plasma; DB: diamond bur; SC: air abrasion with silica-coated $\text{Al}_2\text{O}_3$ particles; FRC: flowable resin composite; LA: laser

SB+FRC (P-score = 0.90) had the greatest likelihood of being the optimal surface treatment for increasing (micro)tensile bond strength, trailed by SC+SI+total (P-score = 0.82) and DB+SB+SI+total (P-score = 0.81). Ultimately, the two interventions with the greatest likelihood of yielding higher (micro)shear and (micro)tensile bond strengths were DB+SI+total (shear MD 41.12 MPa, 95% CI 29.02 to 53.21, P-score 0.99; tensile MD 29.36 MPa, 95% CI 12.36 to 46.35; P-score 0.69), and SC+SI+total (shear MD 16.29 MPa, 95% CI 6.54 to 26.05, P-score 0.67; tensile MD 33.86 MPa, 95% CI 16.17 to 51.54; P-score 0.82).

Per the Egger’s test, there was no publication bias in the included studies of (micro)tensile (P=0.14) and (micro)shear (P=0.85) networks (Supplementary material 7).

Evaluation of node-split inconsistency (Supplementary material 8) revealed that only 3 out of 55 indirect network estimates in the (micro)shear network, and 9 of 64 indirect network estimates in the (micro)tensile network differed from their related direct estimates.

## Discussion

The yearly rate of failure of composite resin restorations ranges between 1% and 4% [49-51]. Recent advancements in adhesive dentistry have made repairing partly defective old composite resin restorations a viable option to complete replacement which is accompanied by a number of complications [9]. Therefore, this systematic review and network meta-analysis sought to explore the influence of common surface treatments on the repair bond strength of aged composite resin restorations, rank these surface treatments, and compare them.

Valente et al. [34] conducted the most recent systematic review and meta-analysis investigating the influence of various surface treatments on the repair bond strength of aged composite resin restorations in 2016. Most of their findings were derived from relatively short-term static water storage aging. Consequently, their findings could not accurately represent the long-term repair bond characteristics.

The majority of studies have investigated early or 24-hour bond strength whereas only rigorous aging methods can properly imitate the inherent conditions of oral cavity such as exposure to chemical agents of food, saliva, occlusal forces, and temperature and pH changes [52]. Moreover, most of the studies have not performed secondary aging (after repairing the old composite), whereas the clinical relevance and long-term durability of bond are heavily reliant on the bonded interface which should also be tested under strict aging methods [33].

Some suggest that the 500 cycles of thermocycling recommended by the ISO TR 11450 standard (2003) [53] is insufficient [33] and also that approximately 100,000 cycles [54] ] or three months of storing in water [55] are required for an appropriate similarity to in-vivo aging and significant bond degradation occurrence. However, due to the lack of studies imposing such strict aging on composite resins, the aging limitation was determined to be at least 1 month or 5000 cycles in the current study based on a similar systematic review [56] which might be closer to the oral environment.

In comparison with the control, the diamond bur + silane + total-etch adhesive and air abrasion with silica-coated Al_2_O_3_ particles + silane + total-etch adhesive were the most effective methods for producing high (micro)tensile and (micro)shear bond strengths (Table 1). In addition, they did not significantly differ with the cohesive strength (Figure 4 and Supplementary material 6), making these pretreatments suitable.

Over 80 percent of dentists are reportedly inclined to utilize identical surface treatment for composite repair as they use for restoration replacement, which is a roughening with diamond bur, accompanied by acid etching and adhesive, respectively [57]. Consequently, the first method looks to be the most practical due to its availability, convenience and lower cost with no need of additional equipment in dental practice.

Silica-coated alumina particles not just roughen the surface of old composite resins but also end up leaving behind a silica-rich layer that promotes bonding by chemical coupling with the following silane [34]. Silanization also improves surface wettability; nevertheless, its influence on chemical coupling improvement is dependent on the presence of silica on the old composite’s surface, that can be provided by silica-coated alumina particles or the structure of composite resins. This is improbable. Because the deterioration of composite resin structure caused by imposed aging breaks the filler-polymer bond, leading to the erosion of surface glass particles [14].

In the (micro)shear network, the second-best technique was surface roughening by lasers followed by silane and total-etch adhesives. This technique was recently presented in in-vitro studies. Neither of the included articles examined the laser’s efficacy as a measure of (micro)shear bond strength. Therefore, more research to investigate the effect of lasers as surface treatments on the repair bond strength of properly aged composite resins is expected.

As per league tables (Supplementary material 6), physical surface treatments alone did not affect the (micro)tensile strength compared to the control (abrasion paper); nevertheless, a substantial difference was found in favor of air abrasion with alumina on its own or silica-coated alumina in the (micro)shear network.

In addition, there have been no distinctions between total-etch adhesives and self-etch adhesives. Also, a chemical treatment is required alongside the mechanical treatment according to tensile network (Supplementary material 6). Utilizing a chemical substance such as silane or adhesive resulted in a (micro)tensile network with a significantly improved bond strength. Eleven comparisons revealed its efficacy, while three studies revealed no distinction (Supplementary material 6). This is consistent with Valente et al. [34] Owing to the (micro)shear network’s substantial inconsistency (I^2^ = 94.1), the findings of these two kinds of adhesive were ambiguous.

The use of phosphoric acid has a positive impact on enamel and dentin adhesion; it does not influence the shear bond strength of the repaired composite resin, since phosphoric acid is too weak to create sufficient surface irregularities. Instead, the monomer components of adhesive systems, such as MDP, has a considerable impact on bond strength [42].

Per the given league tables (Supplementary material 6), there was no significant distinction between comparisons (ten comparisons) with or without the use of silane coupling agents, except for dental burs (two comparisons), for which silane-based surface treatments produced stronger bond strength. A silane coupling agent is used to improve the surface wettability of the composite substrate surface. It also creates covalent bonds with the exposed filler particles on the surface of the old composite resin and copolymerizes with the methacrylate groups of the resin repair material. In the absence of other chemical agents, silanizing the surface of a composite that has been roughened with a diamond bur may strengthen the bond strength. Nevertheless, using silane along with the adhesive agents may have minimal impact on the bond strength [25]. This is consistent with 2 meta-analyses reported previously [34, 38].

Note that most head-to-head assessments through both networks consisted of a single trial. Furthermore, the global heterogeneity in the networks was moderate (I^2^ = 56.9 in the (micro)tensile network) to high (I^2^ = 94.1 in the (micro)shear network).

Calculating sample size seemed more arbitrary than via power analyses, which might compromise the external validity of the results of the investigations. In addition, standard preparation of samples by a single operator and selection of standard specimens (assessment of the prepared samples for the presence of microcracks, voids, and deformities) by direct observation or using microscopes, were not mentioned in over 80 percent of the included studies. Also, in neither of the experiments was the evaluating machine operator blinded.

Regrettably, poor adherence to the aforementioned areas of bias is prevalent in lab experiments, which significantly impacts the pooled results and should be considered in future studies. Ultimately, for clinical application, these results should be taken with caution.

### Strengths and Limitations

The advantages of the present research include providing a network meta-analysis that ranked the widely used clinical surface treatments in terms of being best to yield strong repair bond strength of aged composite resin restorations, comparing these surface treatments despite the lack of direct comparisons, including more rigorous aging methods both before and after repairing composite resins, and attempting to cover more databases than the earlier conducted meta-analysis [34].

Nonetheless, it had certain limitations. This research summarized the existing evidence from laboratory experiments, which may not provide identical results under oral conditions. Yet, they are the primary investigations of clinical tests, and their connection with clinical outcomes is satisfactory [33]. In addition, the limited number of included studies led to the majority of comparisons in the networks being derived from single trials; hence, this problem may have influenced the results.

The focus of the present investigation was on the durability of the repair bond. Other features of the bonded interface, including micro-leakage, marginal adaptation (gap formation), and sealing ability, must also be examined in light of various surface treatment procedures.

## Conclusions

Finally, two network meta-analyses presented that the most effective methods to increase (micro)tensile and (micro)shear bond strength were diamond bur + silane + total-etch adhesives and silica-coated Al_2_O_3_ particles + silane + total-etch adhesive. Because of the first method’s availability, relatively inexpensive, and minimum equipment requirements, it can be used to repair existing composite resin restorations. Research that is well-designed and free of bias should be conducted in future.

## Supporting information

Supplementary material 1. PRISMA checklist for reporting network meta-analysis.

Supplementary material 2. Search strategies in the databases.

Supplementary material 3. List of excluded articles with reasons.

Supplementary material 4. Data extraction table.

Supplementary material 5. Risk of bias assessment of individual studies.

Supplementary material 6. League tables (two-by-two comparisons).

Supplementary material 7. Publication bias (funnel plots).

Supplementary material 8. Node-split inconsistency results.

## Data Availability

The data used to support the findings of this study are available from the corresponding author upon request.

## Supplementary Materials

**Supplementary material 1**. PRISMA checklist for reporting network meta-analysis.

**Supplementary material 2**. Search strategies in the databases.

**Supplementary material 3**. List of excluded articles with reasons.

**Supplementary material 4**. Data extraction table.

**Supplementary material 5**. Risk of bias assessment of individual studies.

**Supplementary material 6**. League tables (two-by-two comparisons).

**Supplementary material 7**. Publication bias (funnel plots).

**Supplementary material 8**. Node-split inconsistency results.

