## Supplementary material 2. Search strategies in the databases. for "Which surface treatment improves the long-term repair bond strength of aged methacrylate-based composite resin restorations? A systematic review and network meta-analysis"

**Supplementary file 2. Search strategies in the databases.**

**PUBMED**

(((repair*[Text Word]) OR (reparation[Text Word])) AND ((((("Composite Resins"[MeSH Terms]) OR (Composite Resin*[Text Word])) OR (Resin, Composit*[Text Word])) OR ("Bisphenol A-Glycidyl Methacrylate"[Mesh])) OR (methacrylic resin*[Text Word]))) AND ("Dentistry"[Mesh] OR dent*)

**Embase**

('resin'/exp OR 'composite resin*':ti,ab OR 'resin, composit*':ti,ab OR 'bisphenol a-glycidyl methacrylate':ti,ab OR 'methacrylic resin*':ti,ab) AND ('repair*':ti,ab OR 'reparation':ti,ab) AND ('dentistry'/exp OR 'dentistry' OR 'dent*' OR 'dent*':ti,ab)

**Scopus**

( TITLE-ABS-KEY ( "resin*" OR "Composite Resin*" OR "Resin Composit*" OR {Bisphenol A Glycidyl Methacrylate} OR "methacrylic resin*" ) ) AND ( TITLE-ABS-KEY ( "repair*" OR "reparation" ) ) AND ( LIMIT-TO ( SUBJAREA , "DENT" ) )

**Web of Science**

---------------------------------------------------------------#1--------------------------------------------------------------------------
TS=("resin*" OR "Composite Resin*" OR "Resin Composit*" OR "Bisphenol A Glycidyl Methacrylate" OR "methacrylic resin*")

---------------------------------------------------------------#2--------------------------------------------------------------------------

TS=("repair*" OR "reparation")

---------------------------------------------------------------#3--------------------------------------------------------------------------

TS=('dent*')

---------------------------------------------------------------#3--------------------------------------------------------------------------

#3 AND #2 AND #1
