## Supplementary material 3. List of excluded articles with reasons. for "Which surface treatment improves the long-term repair bond strength of aged methacrylate-based composite resin restorations? A systematic review and network meta-analysis"

| **Supplementary file 3. List of excluded articles with reasons.** | **Reason for exclusion** |
| --- | --- |
| Acharya, G. S. and M. K. Manjunath (2012). "The effect of surface treatments and bonding regimens on microtensile bond strengths of repaired composite: An in vitro study." Journal of Conservative Dentistry **15**(4): 383-387. | Not enough primary aging |
| Ahmadizenouz, G., et al. (2016). "Effect of different surface treatments on the shear bond strength of nanofilled composite repairs." J Dent Res Dent Clin Dent Prospects **10**(1): 9-16. | Not enough primary aging |
| Akgül, S., et al. (2021). "Repair potential of a bulk-fill resin composite: Effect of different surface-treatment protocols." European Journal of Oral Sciences. | Not enough secondary aging |
| Al-Asmar, A. A., et al. (2017). "Evaluating Various Preparation Protocols on the Shear Bond Strength of Repaired Composite." The journal of contemporary dental practice **18**(3): 182-187. | Not enough secondary aging |
| Al-Asmar, A. A., et al. (2018). "Shear bond strength of repaired composite using single bond adhesive." Jordan Medical Journal **52**(1): 49-57. | No secondary aging |
| Alfawaz, Y. F. and M. S. Bin-Shuwaish (2016). "Microtensile bond strength of repaired composite and compomer dental materials using manufacturers’ recommended protocols." Journal of Biomaterials and Tissue Engineering **6**(2): 149-157. | Inappropriate comparisons |
| Alkatheeri, M. S., et al. (2020). "MICROTENSILE BOND STRENGTH EVALUATION OF RESIN COMPOSITE REPAIRED USING UNIVERSAL ADHESIVES." International Journal of Medical Dentistry **24**(1): 9-15. | Not enough secondary aging |
| Alqarni, D., et al. (2019). "The repair bond strength to resin matrix in cured resin composites after water aging." Dental Materials Journal **38**(2): 233-240. | Not enough secondary aging |
| Altinci, P., et al. (2018). "Repair bond strength of nanohybrid composite resins with a universal adhesive." Acta Biomater Odontol Scand **4**(1): 10-19. | Not enough primary aging |
| Andrade, A. P., et al. (2017). "Composite resin repairs: What is the most effective protocol?" Brazilian Dental Science **20**(1): 99-109. | Not enough secondary aging |
| Aquino, C., et al. (2020). "Repair Bond Strength and Leakage of Non-Aged and Aged Bulk-fill Composite." Oral health & preventive dentistry **18**(4): 783-791. | Not enough secondary aging |
| Arslan Aydoğan, C. and D. Eren (2018). "Effect of different surface treatments and composite resins used for repairing nanohybrid resins on microleakege." Cumhuriyet Dental Journal **21**(3): 184-194. | Not in English |
| Ayar, M. K., et al. (2019). "Repair of aged bulk-fill composite with posterior composite: Effect of different surface treatments." Journal of esthetic and restorative dentistry : official publication of the American Academy of Esthetic Dentistry ... [et al.] **31**(3): 246-252. | Not enough primary aging |
| Bacchi, A., et al. (2013). "Repair bond strength in aged methacrylate-and silorane-based composites." Journal of Adhesive Dentistry **15**(5): 447-452. | Not enough secondary aging |
| Baena, E., et al. (2015). "Influence of repair procedure on composite-to-composite microtensile bond strength." American Journal of Dentistry **28**(5): 255-260. | Not enough primary aging |
| Batista, G. R., et al. (2015). "Effects of different surface treatments on composite repairs." Journal of Adhesive Dentistry **17**(5): 421-426. | Not enough secondary aging |
| Baur, V. and N. Ilie (2013). "Repair of dental resin-based composites." Clinical Oral Investigations **17**(2): 601-608. | Inappropriate comparisons |
| Bektas, Ö. Ö., et al. (2012). "Effect of thermocycling on the bond strength of composite resin to bur and laser treated composite resin." Lasers in Medical Science **27**(4): 723-728. | Not enough secondary aging |
| Blum, I. R., et al. (2021). "Effects of different surface treatments and adhesive self-etch functional monomers on the repair of bulk fill composites: A randomised controlled study." Journal of Dentistry **108**: 103637. | Not enough secondary aging |
| Bonstein, T., et al. (2005). "Evaluation of varied repair protocols applied to aged composite resin." Journal of Adhesive Dentistry **7**(1): 41-49. | Not enough primary aging |
| Bouschlicher, M. R., et al. (1997). "Surface treatment techniques for resin composite repair." American Journal of Dentistry **10**(6): 279-283. | Not enough primary aging |
| Boyer, D. B., et al. (1984). "Build-up and repair of light-cured composites: bond strength." Journal of Dental Research **63**(10): 1241-1244. | Not enough primary aging |
| Brendeke, J. and M. Özcan (2007). "Effect of physicochemical aging conditions on the composite-composite repair bond strength." Journal of Adhesive Dentistry **9**(4): 399-406. | No secondary aging |
| Brosh, T., et al. (1997). "Effect of combinations of surface treatments and bonding agents on the bond strength of repaired composites." The Journal of Prosthetic Dentistry **77**(2): 122-126. | Not enough primary aging |
| Brum, R. T., et al. (2017). "Effect of organic solvents compared to sandblasting on the repair bond strength of nanohybrid and nanofilled composite resins." Indian journal of dental research : official publication of Indian Society for Dental Research **28**(4): 433-441. | Not enough secondary aging |
| Burnett, L. H., et al. (2004). "Tensile Bond Strength of a One-Bottle Adhesive System to Indirect Composites Treated with Er:YAG Laser, Air Abrasion, or Fluoridric Acid." Photomedicine and Laser Surgery **22**(4): 351-U395. | No primary aging |
| Çakir, N. N., et al. (2018). "Bonding performance of universal adhesives on composite repairs, with or without silane application." Journal of Conservative Dentistry **21**(3): 263-268. | Not enough primary aging |
| Causton, B. E. (1975). "Repair of abraded composite fillings. An in vitro study." British Dental Journal **139**(7): 286-288. | Full-text not available (No full text before 1999 in this journal) |
| Cavalcanti, A. N., et al. (2007). "Effect of surface treatments and bonding agents on the bond strength of repaired composites." Journal of Esthetic and Restorative Dentistry **19**(2): 90-98. | Not enough primary aging |
| Celik, E. U., et al. (2011). "Tensile bond strength of an aged resin composite repaired with different protocols." The journal of adhesive dentistry **13**(4): 359-366. | Not enough secondary aging |
| Chan, K. C. and D. B. Boyer (1983). "Repair of conventional and microfilled composite resins." The Journal of Prosthetic Dentistry **50**(3): 345-350. | Not enough primary aging |
| Cho, S. D., et al. (2013). "Effect of Er,Cr:YSGG laser, air abrasion, and silane application on repaired shear bond strength of composites." Operative Dentistry **38**(3): E1-9. | Not enough secondary aging |
| Consani, R. L. X., et al. (2016). "Repair strength in simulated restorations of methacrylate- or silorane-based composite resins." Brazilian Dental Journal **27**(4): 463-467. | Inappropriate comparisons |
| Costa, T. R. F., et al. (2010). "Durability of surface treatments and intermediate agents used for repair of a polished composite." Operative Dentistry **35**(2): 231-237. | Not enough primary aging |
| Crumpler, D. C., et al. (1989). "Bonding to resurfaced posterior composites." Dental materials : official publication of the Academy of Dental Materials **5**(6): 417-424. | Not enough primary aging |
| Cuevas-Suárez, C. E., et al. (2020). "Repair bond strength of bulk-fill resin composite: Effect of different adhesivprotocols." Dental Materials Journal **39**(2): 236-241. | Not enough secondary aging |
| Cura, M., et al. (2016). "Effect of surface treatment and aging on bond strength of composite resin onlays." Journal of Prosthetic Dentistry **116**(3): 389-396. | No primary aging |
| Da Costa, T. R. F., et al. (2012). "Durability of composite repair using different surface treatments." Journal of Dentistry **40**(6): 513-521. | Not enough primary aging |
| da Silva, C. L., et al. (2020). "Does use of silane-containing universal adhesive eliminate the need for silane application in direct composite repair?" Brazilian Oral Research **34**. | Not enough secondary aging |
| Dall'oca, S., et al. (2008). "Repair potential of a laboratory-processed nano-hybrid resin composite." Journal of Oral Science **50**(4): 403-412. | Not enough primary aging |
| De Jesus Tavarez, R. R., et al. (2017). "Shear bond strength of different surface treatments in bulk fill, microhybrid, and nanoparticle repair resins." Clinical, Cosmetic and Investigational Dentistry **9**: 61-66. | Not enough primary aging |
| de Medeiros, T. C., et al. (2019). "Repair bond strength of bulk fill composites after different adhesion protocols." J Clin Exp Dent **11**(11): e1000-e1005. | Not enough secondary aging |
| de Melo, M. A. V., et al. (2011). "Effects of different surface treatments and accelerated artificial aging on the bond strength of composite resin repairs." Brazilian Oral Research **25**(6): 485-491. | Not enough secondary aging |
| Dhuru, V. B. and C. H. Lloyd (1986). "The fracture toughness of repaired composite." Journal of Oral Rehabilitation **13**(5): 413-421. | Not enough primary aging |
| Dias, A. A. M., et al. (2011). "Flexural bond strength of repaired composite resin restorations: Influence of surface treatments and aging." General Dentistry **59**(2): e82-e86. | Not enough secondary aging |
| Dias, W. R. L., et al. (2003). "Repairability of a packable resin-based composite using different adhesives." American Journal of Dentistry **16**(3): 181-185. | Not enough primary aging |
| Duran, İ., et al. (2015). "Effects of Er:YAG Laser Pretreatment with Different Energy Levels on Bond Strength of Repairing Composite Materials." Photomedicine and Laser Surgery **33**(6): 320-325. | Not enough secondary aging |
| Dursun, M. N., et al. (2021). "The effect of different surface preparation methods and various aging periods on microtensile bond strength for composite resin repair." Nigerian Journal of Clinical Practice **24**(2): 282-291. | Not enough secondary aging |
| Eren, D., et al. (2013). "Can Er: YAG laser be an alternative to conventional methods for repairing composite resins?" Cumhuriyet Dental Journal **16**(2): 125-132. | Not enough primary aging |
| Eren, D., et al. (2019). "Effect of Different Surface Treatments and Roughness on the Repair Bond Strength of Aged Nanohybrid Composite." Photobiomodulation Photomedicine and Laser Surgery **37**(8): 473-482. | Not enough secondary aging |
| Fawzy, A. S., et al. (2008). "Effect of surface treatments on the tensile bond strength of repaired water-aged anterior restorative micro-fine hybrid resin composite." Journal of Dentistry **36**(12): 969-976. | Not enough secondary aging |
| Fornazari, I. A., et al. (2020). "Reliability and correlation between microshear and microtensile bond strength tests of composite repairs." Journal of the Mechanical Behavior of Biomedical Materials **103**. | Not enough secondary aging |
| Fornazari, I. A., et al. (2017). "Effect of Surface Treatment, Silane, and Universal Adhesive on Microshear Bond Strength of Nanofilled Composite Repairs." Operative Dentistry **42**(4): 367-374. | Not enough primary aging |
| Frankenberger, R., et al. (2003). "Fatigue behavior of the resin-resin bond of partially replaced resin-based composite restorations." American Journal of Dentistry **16**(1): 17-22. | Not enough secondary aging |
| Ghavam, M., et al. (2018). "Repair bond strength of composite: Effect of surface treatment and type of composite." Journal of Clinical and Experimental Dentistry **10**(6): e520-e527. | Not enough secondary aging |
| Goncalves, A. P., et al. (2015). "Short Exposure to 1% Hydrofluoric Acid to Improve the Repair Bond Strength of Dental Resin Composites." Journal of Adhesion **91**(3): 235-243. | Not enough primary aging |
| Gonulol, N., et al. (2019). "Shear Bond Strength of Aged Composite Restorations Repaired with a Universal Injectable Composite." European Journal of Therapeutics **25**(4): 273-278. | Not enough primary aging |
| Gregory, W. A., et al. (1992). "Physical properties and repair bond strength of direct and indirect composite resins." The Journal of Prosthetic Dentistry **68**(3): 406-411. | Not enough primary aging |
| Gregory, W. A., et al. (1990). "Bond strengths of chemically dissimilar repaired composite resins." The Journal of Prosthetic Dentistry **64**(6): 664-668. | Not enough primary aging |
| Gupta, S., et al. (2015). "A comparative effect of various surface chemical treatments on the resin composite-composite repair bond strength." Journal of Indian Society of Pedodontics and Preventive Dentistry **33**(3): 245-249. | Not enough primary aging |
| Gutierrez, N. C., et al. (2019). "Bond strength of composite resin restoration repair: Influence of silane and adhesive systems." Journal of Contemporary Dental Practice **20**(8): 880-886. | No secondary aging |
| Hagge, M. S., et al. (2002). "Shear bond strength of bis-acryl composite provisional material repaired with flowable composite." Journal of esthetic and restorative dentistry : official publication of the American Academy of Esthetic Dentistry ... [et al.] **14**(1): 47-52. | Not enough primary aging |
| Hamano, N., et al. (2011). "Effect of different surface treatments on the repair strength of a nanofilled resin-based composite." Dental Materials Journal **30**(4): 537-545. | Not enough primary aging |
| Hannig, C., et al. (2006). "Shear bond strength of repaired adhesive filling materials using different repair procedures." Journal of Adhesive Dentistry **8**(1): 35-40. | Inappropriate comparisons |
| Hemadri, M., et al. (2014). "Shear Bond Strength of Repaired Composites Using Surface Treatments and Repair Materials: An In vitro Study." J Int Oral Health **6**(6): 22-25. | Not enough primary aging |
| Hisamatsu, N., et al. (2002). "Effect of silane primers and unfilled resin bonding agents on repair bond strength of a prosthodontic microfilled composite." Journal of Oral Rehabilitation **29**(7): 644-648. | No primary aging |
| Ilie, N. and B. Stawarczyk (2015). "Efficiency of different repair kits on bonding to aged dental resin composite substrates." International Journal of Adhesion and Adhesives **58**: 7-12. | No primary aging |
| Imbery, T. A., et al. (2014). "Evaluation of flexural, diametral tensile, and shear bond strength of composite repairs." Operative Dentistry **39**(6): E250-E260. | Not enough primary aging |
| Irmak, O., et al. (2017). "Adhesive system affects repair bond strength of resin composite." J Istanb Univ Fac Dent **51**(3): 25-31. | Not enough secondary aging |
| Ivanovas, S., et al. (2011). "How to repair fillings made by silorane-based composites." Clinical Oral Investigations **15**(6): 915-922. | Not enough primary aging |
| Jafarzadeh Kashi, T. S., et al. (2011). "An in vitro assessment of the effects of three surface treatments on repair bond strength of aged composites." Oper Dent **36**(6): 608-617. | Not enough primary aging |
| Joulaei, M., et al. (2012). "Effect of Different Surface Treatments on Repair Micro-shear Bond Strength of Silica- and Zirconia-filled Composite Resins." J Dent Res Dent Clin Dent Prospects **6**(4): 131-137. | Not enough secondary aging |
| Junior, S. A. R., et al. (2009). "Influence of surface treatments on the bond strength of repaired resin composite restorative materials." Dental Materials **25**(4): 442-451. | Not enough primary aging |
| Kashi, T. S. J., et al. (2011). "An in vitro assessment of the effects of three surface treatments on repair bond strength of aged composites." Operative Dentistry **36**(6): 608-617. | Not enough primary aging |
| Kimyai, S., et al. (2010). "Comparison of the effect of three mechanical surface treatments on the repair bond strength of a laboratory composite." Photomedicine and Laser Surgery **28 Suppl 2**: S25-30. | Not enough primary aging |
| Kimyai, S., et al. (2015). "Effect of different mechanical and chemical surface treatments on the repaired bond strength of an indirect composite resin." Lasers in Medical Science **30**(2): 653-659. | Not enough primary aging |
| Koç-Vural, U., et al. (2017). "Bond strength of dental nanocomposites repaired with a bulkfill composite." Journal of Clinical and Experimental Dentistry **9**(3): e437-e442. | Inappropriate comparisons |
| Kupiec, K. A. and W. W. Barkmeier (1996). "Laboratory evaluation of surface treatments for composite repair." Operative Dentistry **21**(2): 59-62. | No primary aging |
| Lemos, C. A., et al. (2016). "Repairability of aged resin composites mediated by different restorative systems." Acta odontologica latinoamericana : AOL **29**(1): 7-13. | No secondary aging |
| Lewis, G., et al. (1998). "Shear bond strength of immediately repaired light-cured composite resin restorations." Operative Dentistry **23**(3): 121-127. | No primary aging |
| Lin, F., et al. (2015). "Microtensile strength of composite-composite bonding: an in vitro study." Beijing da xue xue bao. Yi xue ban = Journal of Peking University. Health sciences **47**(1): 124-128. | Not enough primary aging |
| Liu, C., et al. (2015). "Effect of thermal cycling on the composite- composite repair bond strength." Zhonghua kou qiang yi xue za zhi = Zhonghua kouqiang yixue zazhi = Chinese journal of stomatology **50**(8): 483-487. | Not in English |
| Lloyd, C. H. and V. B. Dhuru (1985). "Effect of a commercial bonding agent upon the fracture toughness (K'IC) of repaired heavily filled composite." Dental materials : official publication of the Academy of Dental Materials **1**(3): 83-85. | Not enough primary aging |
| Loomans, B. A. C., et al. (2017). "Effect of different surface treatment techniques on the repair strength of indirect composites." Journal of Dentistry **59**: 18-25. | No primary aging |
| Loomans, B. A. C., et al. (2011). "Is there one optimal repair technique for all composites?" Dental Materials **27**(7): 701-709. | Not enough secondary aging |
| Lucena-Martín, C., et al. (2001). "The effect of various surface treatments and bonding agents on the repaired strength of heat-treated composites." The Journal of Prosthetic Dentistry **86**(5): 481-488. | Not enough secondary aging |
| Magni, E., et al. (2011). "Influence of ozone on the composite-to-composite bond." Clinical Oral Investigations **15**(2): 249-256. | Not enough primary aging |
| Maneenut, C., et al. (2011). "The repair potential of resin composite materials." Dental materials : official publication of the Academy of Dental Materials **27**(2): e20-27. | Not enough secondary aging |
| Mirzaie, M., et al. (2016). "Surface treatment comparison by application of diamond bur and Er,Cr:YSGG at different powers: Morphological and mechanical evaluation." Laser Therapy **25**(3): 215-220. | No primary aging |
| Mitsaki-Matsou, H., et al. (1991). "An in vitro study of the tensile strength of composite resins repaired with the same or another composite resin." Quintessence international (Berlin, Germany : 1985) **22**(6): 475-481. | No secondary aging |
| Moncada, G., et al. (2012). "Bond strength evaluation of nanohybrid resin-based composite repair." General Dentistry **60**(3): 230-234. | Not enough primary aging |
| Mossa, H., et al. (2016). "Effect of hyperbaric oxygen profiles on the bond strength of repaired composite resin." Journal of International Society of Preventive and Community Dentistry **6**: S70-S74. | Not enough primary aging |
| Nagano, D., et al. (2018). "Effect of water aging of adherend composite on repair bond strength of nanofilled composites." Journal of Adhesive Dentistry **20**(5): 425-433. | Not enough secondary aging |
| Nassoohi, N., et al. (2015). "Effects of three surface conditioning techniques on repair bond strength of nanohybrid and nanofilled composites." Dental Research Journal **12**(6): 554-561. | Not enough secondary aging |
| Oh, H. K. and D. H. Shin (2021). "Effect of adhesive application method on repair bond strength of composite." Restor Dent Endod **46**(3): e32. | Not enough secondary aging |
| Oliveira, P. H. C., et al. (2019). "Effect of Surface Treatment with CO2 Laser on Bond Strength in Composite Resin Restorations." Photobiomodulation Photomedicine and Laser Surgery **37**(7): 428-433. | Not enough primary aging |
| Özcan, M., et al. (2005). "Effect of three surface conditioning methods to improve bond strength of particulate filler resin composites." Journal of Materials Science: Materials in Medicine **16**(1): 21-27. | No primary aging |
| Özcan, M., et al. (2007). "Effect of surface conditioning methods on the microtensile bond strength of resin composite to composite after aging conditions." Dental Materials **23**(10): 1276-1282. | Not enough secondary aging |
| Özcan, M., et al. (2013). "Repair bond strength of microhybrid, nanohybrid and nanofilled resin composites: Effect of substrate resin type, surface conditioning and ageing." Clinical Oral Investigations **17**(7): 1751-1758. | No secondary aging |
| Ozcan, M., et al. (2010). "Effect Aging Conditions on the Repair Bond Strength of a Microhybrid and a Nanohybrid Resin Composite." Journal of Adhesive Dentistry **12**(6): 451-459. | No secondary aging |
| Ozcan, M., et al. (2014). "Adhesion of substrate-adherent combinations for early composite repairs: Effect of intermediate adhesive resin application." International Journal of Adhesion and Adhesives **49**: 97-102. | No primary aging |
| Özel Bektas, Ö., et al. (2012). "Effect of thermocycling on the bond strength of composite resin to bur and laser treated composite resin." Lasers Med Sci **27**(4): 723-728. | Not enough secondary aging |
| Oztas, N., et al. (2003). "The effect of air abrasion with two new bonding agents on composite repair." Operative Dentistry **28**(2): 149-154. | Not enough primary aging |
| Padipatvuthikul, P. and L. H. Mair (2007). "Bonding of composite to water aged composite with surface treatments." Dental Materials **23**(4): 519-525. | Not enough secondary aging |
| Papacchini, F., et al. (2007). "Composite-to-composite microtensile bond strength in the repair of a microfilled hybrid resin: Effect of surface treatment and oxygen inhibition." Journal of Adhesive Dentistry **9**(1): 25-31. | Not enough primary aging |
| Papacchini, F., et al. (2007). "Effect of intermediate agents and pre-heating of repairing resin on composite-repair bonds." Oper Dent **32**(4): 363-371. | Not enough secondary aging |
| Papacchini, F., et al. (2007). "Effect of air-drying temperature on the effectiveness of silane primers and coupling blends in the repair of a microhybrid resin composite." Journal of Adhesive Dentistry **9**(4): 391-397. | Not enough secondary aging |
| Papacchini, F., et al. (2007). "The application of hydrogen peroxide in composite repair." Journal of Biomedical Materials Research - Part B Applied Biomaterials **82**(2): 298-304. | Not enough secondary aging |
| Papacchini, F., et al. (2008). "Flowable composites as intermediate agents without adhesive application in resin composite repair." American Journal of Dentistry **21**(1): 53-58. | Not enough secondary aging |
| Passos, S. P., et al. (2007). "Bond strength durability of direct and indirect composite systems following surface conditioning for repair." Journal of Adhesive Dentistry **9**(5): 443-447. | No primary aging |
| Perriard, J., et al. (2009). "The Effect of Water Storage, Elapsed Time and Contaminants on the Bond Strength and Interfacial Polymerization of a Nanohybrid Composite." Journal of Adhesive Dentistry **11**(6): 469-478. | Not enough secondary aging |
| Pinheiro, I., et al. (2019). "Effect of surface treatment and the use of mouthwashes on repaired composite bond strength." Revista Portuguesa de Estomatologia, Medicina Dentaria e Cirurgia Maxilofacial **60**(3): 130-136. | Not enough primary aging |
| Pontes, A. P., et al. (2005). "Shear bond strength of direct composite repairs in indirect composite systems." General Dentistry **53**(5): 343-347. | Not enough secondary aging |
| Puleio, F., et al. (2020). "Chemical and Mechanical Roughening Treatments of a Supra-Nano Composite Resin Surface: SEM and Topographic Analysis." Applied Sciences-Basel **10**(13). | Inappropriate comparisons |
| Rathke, A., et al. (2009). "Effect of different surface treatments on the composite-composite repair bond strength." Clinical Oral Investigations **13**(3): 317-323. | Not enough secondary aging |
| Ribeiro, J. C. R., et al. (2008). "Shear strength evaluation of composite-composite resin associations." Journal of Dentistry **36**(5): 326-330. | No primary aging |
| Rinastiti, M., et al. (2011). "Effects of surface conditioning on repair bond strengths of non-aged and aged microhybrid, nanohybrid, and nanofilled composite resins." Clinical Oral Investigations **15**(5): 625-633. | No secondary aging |
| Ritter, A. V., et al. (2020). "Effect of Tribochemical Coating on Composite Repair Strength." Operative Dentistry **45**(6): E334-E342. | Not enough secondary aging |
| Ritter, A. V., et al. (2019). "Composite-composite Adhesion as a Function of Adhesive-composite Material and Surface Treatment." Operative Dentistry **44**(4): 348-354. | Not enough secondary aging |
| Rodrigues, S. A., Jr., et al. (2009). "Influence of surface treatments on the bond strength of repaired resin composite restorative materials." Dent Mater **25**(4): 442-451. | Not enough primary aging |
| Rossato, D. M., et al. (2009). "Influence of Er:YAG laser on surface treatment of aged composite resin to repair restoration." Laser Physics **19**(11): 2144-2149. | Not enough primary aging |
| Sadaghiani, M., et al. (2018). "Comparison of the Micro-Tensile Bond Strength of Composite Resin Restoration in Micro- and Nano-hybrid Composite Resins using Different Interfacial Materials." Journal of Research in Medical and Dental Science **6**(3): 436-444. | Not enough secondary aging |
| Saunders, W. P. (1990). "Effect of fatigue upon the interfacial bond strength of repaired composite resins." Journal of Dentistry **18**(3): 158-162. | Not enough primary aging |
| Shafiei, F., et al. (2017). "Effect of timing of repair-on-repair bond strength of methacrylate- and silorane-based composite resins." General Dentistry **65**(3): 45-49. | Not enough secondary aging |
| Shahdad, S. A. and J. G. Kennedy (1998). "Bond strength of repaired anterior composite resins: an in vitro study." Journal of Dentistry **26**(8): 685-694. | Not enough primary aging |
| Shinohara, A., et al. (2018). "Effects of three silane primers and five adhesive agents on the bond strength of composite material for a computer-aided design and manufacturing system." Journal of applied oral science : revista FOB **26**: e20170342. | No primary aging |
| Silva, C. L. D., et al. (2020). "Does use of silane-containing universal adhesive eliminate the need for silane application in direct composite repair?" Brazilian Oral Research **34**: e045. | Not enough secondary aging |
| Sismanoglu, S. (2019). "Efficiency of self-adhering flowable resin composite and different surface treatments in composite repair using a universal adhesive." Nigerian Journal of Clinical Practice **22**(12): 1675-1679. | Not enough secondary aging |
| Şişmanoĝlu, S. (2019). "Effect of different surface treatments on the repair of aged bulk-fill composites: An in vitro study." Cumhuriyet Dental Journal **22**(4): 451-460. | Not enough secondary aging |
| Sismanoglu, S., et al. (2020). "Efficacy of different surface treatments and universal adhesives on the microtensile bond strength of bulk-fill composite repair." Journal of Adhesion Science and Technology **34**(10): 1115-1127. | Not enough secondary aging |
| SÖDerholm, K. J. M. (1986). "Flexure strength of repaired dental composites." European Journal of Oral Sciences **94**(4): 364-369. | Not enough primary aging |
| Sousa, A. B. S., et al. (2013). "Effect of various aging protocols and intermediate agents on the bond strength of repaired composites." Journal of Adhesive Dentistry **15**(2): 137-144. | No secondary aging |
| Souza, E. M., et al. (2008). "Effect of different surface treatments on the repair bond strength of indirect composites." American Journal of Dentistry **21**(2): 93-96. | Not enough primary aging |
| Spyrou, M., et al. (2014). "The reparability of contemporary composite resins." European Journal of Dentistry **8**(3): 353-359. | Not enough primary aging |
| Sullivan, R. H., et al. (2016). "Strengths of additions to composite or resin-modified glass-ionomer." International Journal of Adhesion and Adhesives **69**: 86-90 | No primary aging |
| Swift Jr, E. J., et al. (1994). "Effect of a silane coupling agent on composite repair strengths." American Journal of Dentistry **7**(4): 200-202. | Full-text not available (No full text before 2004 in this journal) |
| Swift Jr, E. J., et al. (1992). "Evaluation of new methods for composite repair." Dental materials : official publication of the Academy of Dental Materials **8**(6): 362-365. | Not enough primary aging |
| Tantbirojn, D., et al. (2015). "Failure Strengths of Composite Additions and Repairs." Operative Dentistry **40**(4): 364-371. | Not enough primary aging |
| Tavarez, R. R. D., et al. (2017). "Shear bond strength of different surface treatments in bulk fill, microhybrid, and nanoparticle repair resins." Clinical Cosmetic and Investigational Dentistry **9**. | Not enough primary aging |
| Teixeira, E. C., et al. (2005). "Shear bond strength of self-etching bonding systems in combination with various composites used for repairing aged composites." Journal of Adhesive Dentistry **7**(2): 159-164. | Not enough secondary aging |
| Tezvergil, A., et al. (2003). "Composite-composite repair bond strength: Effect of different adhesion primers." Journal of Dentistry **31**(8): 521-525. | Not enough primary aging |
| Trajtenberg, C. P. and J. M. Powers (2004). "Bond strengths of repaired laboratory composites using three surface treatments and three primers." American Journal of Dentistry **17**(2): 123-126. | Not enough secondary aging |
| Trajtenberg, C. P. and J. M. Powers (2004). "Effect of hydrofluoric acid on repair bond strength of a laboratory composite." American Journal of Dentistry **17**(3): 173-176. | Not enough primary aging |
| Valente, L. L., et al. (2017). "Repair bond strength of resin composite with experimental primers - effect of formulation variables." Journal of Adhesion Science and Technology **31**(7): 806-815. | Not enough secondary aging |
| Valente, L. L., et al. (2015). "Effect of Diamond Bur Grit Size on Composite Repair." The journal of adhesive dentistry **17**(3): 257-263. | Not enough secondary aging |
| Veiga de Melo, M. A., et al. (2011). "Effects of different surface treatments and accelerated artificial aging on the bond strength of composite resin repairs." Brazilian Oral Research **25**(6): 485-491. | Uncommon aging method (C-UV accelerated aging) |
| Visuttiwattanakorn, P., et al. (2017). "Microtensile bond strength of repaired indirect resin composite." Journal of Advanced Prosthodontics **9**(1): 38-44. | Not enough primary aging |
| Vivas, J., et al. (2009). "Effect of different surface treatments on the shear and flexural re-bond strengths of a micro-hybrid composite." The journal of contemporary dental practice **10**(5): E001-008. | No primary aging |
| Wendler, M., et al. (2016). "Repair Bond Strength of Aged Resin Composite after Different Surface and Bonding Treatments." Materials (Basel) **9**(7). | Not enough secondary aging |
| Yarmohammadi, E. and M. Farshchian (2020). "In Vitro Evaluation of the Effect of Different Surface Treatments on Shear Bond Strength of New to Old Composite Restorations." Dental Hypotheses **11**(4): 108-111. | Not enough primary aging |
| Yesilyurt, C., et al. (2009). "Initial repair bond strength of a nano-filled hybrid resin: Effect of surface treatments and bonding agents." Journal of Esthetic and Restorative Dentistry **21**(4): 251-260. | Not enough secondary aging |
| Staxrud, F. and J. E. Dahl (2015). "Silanising agents promote resin-composite repair." International Dental Journal **65**(6): 311-315. | Mixed silorane and methacrylate-based composites in the aged groups and did not report the outcome separately based on the composite types |
| Pilo, R., et al. (2016). "Effect of Silane Reaction Time on the Repair of a Nanofilled Composite by a Tribochemical Treatment." Journal of Adhesive Dentistry **18**(2): 125-134. | Inappropriate comparisons |
