## Supplementary material 4. Data extraction table. for "Which surface treatment improves the long-term repair bond strength of aged methacrylate-based composite resin restorations? A systematic review and network meta-analysis"

| **Author (Year)** | **Aged composite** | **Repair composite** | **Primary/secondary aging type** | **Group size** | **Surface treatment** | | **Test** | **Bond strength (MPa)** |
| --- | --- | --- | --- | --- | --- | --- | --- | --- |
|  |  |  |  |  | **Mechanical** | **Chemical** |  |  |
| Burrer et al. (2021) | Ceram.x Spectra ST (HV) | Ceram.x Spectra ST (HV) | thermal cycling (5000 cycles, 5–55˚C, dwell time: 20 s, transfer time: 10 s)/same | 8 | G1: abrasive paper | adhesive (OptiBond FL) (total-etch) | µTBS | 19.1 ± 13.0 |
|  |  |  |  |  | G3: abrasive paper + Sandblasting (Al_2_O_3_) | silane + adhesive (OptiBond FL) (total-etch) |  | 37.8 ± 9.5 |
| Karadas (2021) | Charisma Smart | Charisma Smart | thermal cycling (5000 cycles, 5–55˚C, dwell time: 30 s)/ same (30000 cycles) | 60 | G1: abrasive paper | adhesive (All-Bond Universal) (self-etch) | µTBS | 22.99 ± 6.30 |
|  |  |  |  |  | G2: abrasive paper | adhesive (ScotchBond Universal) (self-etch) |  | 29.79 ± 8.01 |
|  |  |  |  |  | G3: abrasive paper | adhesive (Tokuyama Universal Bond) (self-etch) |  | 28.82 ± 10.27 |
|  |  |  |  |  | G4: abrasive paper | No treatment |  | 6.95 ± 2.85 |
| Negreiros et al. (2021) | Filtek Supreme Ultra | Filtek Supreme Ultra  Charisma | 4 months in water (37˚C) /1 year in water (37˚C) | 6 | G1: abrasive paper | No treatment | µSBS | Filtek: 3.6 ± 1.0  Charisma: 1.4 ± 0.3 |
|  |  |  |  |  | G2: abrasive paper + Sandblasting (Al_2_O_3_) | silane + adhesive (Adper Scotchbond Multi-Purpose) (total-etch) |  | Filtek: 20.3 ± 2.9  Charisma: 17.7 ± 1.5 |
|  |  |  |  |  | G3: abrasive paper | adhesive (Adper Scotchbond Multi-Purpose) (total-etch) |  | Filtek: 20.7 ± 4.2  Charisma: 17.8 ± 1.3 |
|  |  |  |  |  | G4: abrasive paper + argon plasma | adhesive (Adper Scotchbond Multi-Purpose) (total-etch) |  | Filtek: 12.8 ± 2.4  Charisma: 14.4 ± 1.4 |
|  |  |  |  |  | G5: abrasive paper + Sandblasting (Al_2_O_3_) | adhesive (Adper Scotchbond Multi-Purpose) (total-etch) |  | Filtek: 20.8 ± 3.5  Charisma: 17.9 ± 1.6 |
|  |  |  |  |  | G6: abrasive paper + Sandblasting (Al_2_O_3_) + argon plasma | adhesive (Adper Scotchbond Multi-Purpose) (total-etch) |  | Filtek: 15.4 ± 5.6  Charisma: 17.3 ± 1.6 |
|  |  |  |  |  | G7: abrasive paper + argon plasma | No treatment |  | Filtek: 3.2 ± 0.6  Charisma: 4.2 ± 1.2 |
| Willers et al. (2021) | Charisma | Charisma | 4 years in water (37˚C)/ 1 year in water (37˚C) | 18 | G1: abrasive paper + sandblasting (Al_2_O_3_) | Silane + adhesive (Adper Scotchbond Multi-Purpose) (total-etch) | SBS | 18.7 ± 3.6 |
|  |  |  |  |  | G2: abrasive paper + sandblasting (Al_2_O_3_) | adhesive (Gluma Bond Universal) (self-etch) |  | 14.3 ± 4.6 |
|  |  |  |  |  | G3: abrasive paper + sandblasting (Al_2_O_3_) | adhesive (Adhese Universal) (self-etch) |  | 18.6 ± 2.9 |
|  |  |  |  |  | G4: abrasive paper + sandblasting (Al_2_O_3_) | adhesive (Scotchbond Bond Universal) (self-etch) |  | 19.8 ± 3.9 |
| Michelotti et al. (2020) | Filtek Supreme XTE | Filtek Supreme XTE | thermal cycling (5000 cycles, 5–55˚C, dwell time: 20 s; transfer time: 10 s; duration of each cycle: 50 s)/same | 6 | G1: abrasive paper + diamond bur | adhesive (Scotchbond Universal) (self-etch) | µTBS | 27.13 ± 1.24 |
|  |  |  |  |  | G2: abrasive paper + diamond bur | silane + adhesive (Scotchbond Universal) (self-etch) |  | 31.62 ± 4.36 |
|  |  |  |  |  | G3: abrasive paper + diamond bur | silane + adhesive (OptiBond FL) (total-etch) |  | 35.43 ± 3.24 |
|  |  |  |  |  | G4: abrasive paper + diamond bur | No treatment |  | 2.94 ± 2.78 |
|  |  |  |  |  | G5: abrasive paper + sandblasting (Al_2_O_3_) | adhesive (Scotchbond Universal) (self-etch) |  | 32.52 ± 5.74 |
|  |  |  |  |  | G6: abrasive paper + sandblasting (Al_2_O_3_) | silane + adhesive (Scotchbond Universal) (self-etch) |  | 37.04 ± 3.03 |
|  |  |  |  |  | G7: abrasive paper + sandblasting (Al_2_O_3_) | silane + adhesive (OptiBond FL) (total-etch) |  | 39.88 ± 5.53 |
|  |  |  |  |  | G8: abrasive paper + sandblasting (Al_2_O_3_) | No treatment |  | 12.37 ± 1.73 |
|  |  |  |  |  | G9: abrasive paper + silica coating | adhesive (Scotchbond Universal) (self-etch) |  | 33.56 ± 0.50 |
|  |  |  |  |  | G10: abrasive paper + silica coating | silane + adhesive (Scotchbond Universal) (self-etch) |  | 35.01 ± 5.09 |
|  |  |  |  |  | G11: abrasive paper + silica coating | silane + adhesive (OptiBond FL) (total-etch) |  | 39.97 ± 6.98 |
|  |  |  |  |  | G12: abrasive paper + silica coating | No treatment |  | 19.90 ± 3.58 |
| Moura et al. (2020) | Filtek Z350 | Filtek Z350 | 6 months in water/  thermal cycling (10000 cycles; 5–55˚C; dwell time: 30 s) | 12 | G1: abrasive paper + silica coating | adhesive (Scotchbond Universal) (self-etch) | SBS | 20.92 ± 7.29 |
|  |  |  |  |  | G2: abrasive paper + sandblasting (Al_2_O_3_) | adhesive (Scotchbond Universal) (self-etch) |  | 18.18 ± 5.6 |
| Dieckmann et al. (2020) | Filtek supreme XTE | Filtek supreme XTE | thermal cycling (5000 cycles, 5–55˚C; dwell time: 20 s; transfer time: 10 s)/same | 6 | G1: abrasive paper | adhesive (OptiBond FL) (total-etch) | µTBS | 0.00 ± 0.00 |
|  |  |  |  |  | G2: abrasive paper + diamond bur | silane + adhesive (OptiBond FL) (total-etch) |  | 22.98 ± 5.89 |
|  |  |  |  |  | G3: abrasive paper + diamond bur + Sandblasting (Al_2_O_3_) | silane + adhesive (OptiBond FL) (total-etch) |  | 28.43 ± 9.86 |
| Ayres et al. (2019) | Charisma | Charisma | 6 months in water (37˚C)/1 year in water (37˚C) | 10 | G1: abrasive paper + sandblasting (Al_2_O_3_) | silane + adhesive (Adper Scotchbond Multi-Purpose) (total-etch) | µSBS | 23.0 ± 3.2 |
|  |  |  |  |  | G2: abrasive paper + sandblasting (Al_2_O_3_) + argon plasma | silane + adhesive (Adper Scotchbond Multi-Purpose) (total-etch) |  | 20.3 ± 4.3 |
|  |  |  |  |  | G3: abrasive paper + argon plasma | silane + adhesive (Adper Scotchbond Multi-Purpose) (total-etch) |  | 15.9 ± 2.1 |
|  |  |  |  |  | G4: abrasive paper + argon plasma | silane |  | 5.1 ± 0.4 |
|  |  |  |  |  | G5: abrasive paper + argon plasma | adhesive (Adper Scotchbond Multi-Purpose) (total-etch) |  | 13.8 ± 3.4 |
|  |  |  |  |  | G6: abrasive paper + argon plasma | No treatment |  | 1.8 ± 0.6 |
| Demirel and Gur (2019) | Clearfil Majesty Esthetic | Clearfil Majesty Esthetic | thermal cycling (10000 cycles; 5–55˚C; dwell time: 30 s; transfer time: 10 s)/same | 11 | G1: abrasive paper + diamond bur | silane + adhesive (Clearfil S3 Bond Plus) (total-etch) | μSBS | 68.85 ± 4.89 |
|  |  |  |  |  | G2: abrasive paper + diamond bur | adhesive (single bond universal) (total-etch) |  | 45.90 ± 6.40 |
|  |  |  |  |  | G3: abrasive paper + diamond bur | adhesive (Clearfil Universal Bond) (total-etch) |  | 48.91 ± 7.90 |
|  |  |  |  |  | G4: abrasive paper + diamond bur | adhesive (Clearfil S3 Bond Plus) (total-etch) |  | 32.36 ± 5.64 |
| Flury et al. (2019) | Filtek Z250 | Filtek Z250 | 3 months in water (37˚C)/1 year in water (37˚C) | 15 | G1: sandblasting (Al_2_O_3_) | silane + adhesive (OptiBond FL) (total-etch) | μSBS | 13.41 ± 3.25 |
|  |  |  |  |  | G2: sandblasting (Al_2_O_3_) | adhesive (Scotchbond Universal) (self-etch) |  | 14.17 ± 2.87 |
| Oglakci and Arhun (2019) | Tetric EvoCeram Bulk Fill | Tetric EvoCeram Bulk Fill  Tetric EvoCeram Nanohybrid | thermal cycling (5000 cycles, 5–55˚C; dwell time: 20 s; transfer time: 10 s)/same | 15 | G1: abrasive disk + diamond bur | adhesive (Tetric N-Bond Universal) (self-etch) | SBS | BF: 24.69 ± 4.82  NH: 20.69 ± 7.17 |
|  |  |  |  |  | G2: abrasive disk + diamond bur | adhesive (Tetric N-Bond Universal) (total-etch) |  | BF: 25.86 ± 5.74  NH: 20.41 ± 3.70 |
|  |  |  |  |  | G3: abrasive disk + diamond bur | adhesive (Clearfil SE Bond) (self-etch) |  | BF: 27.05 ± 4.93  NH: 22.08 ± 6.37 |
|  |  |  |  |  | G4: abrasive disk + diamond bur | adhesive (Adper Single Bond 2) (total-etch) |  | BF: 24.49 ± 6.95  NH: 18.74 ± 6.40 |
| Kouros et al. (2018) | Filtek Ultimate | Filtek Ultimate | 6 months in artificial saliva/thermal cycling (5000 cycles; 5–550˚C; dwell time: 30 s) | 10 | G1: diamond bur | adhesive (Adper Single Bond 2) (total-etch) | SBS | 52.72 ± 10.9 |
|  |  |  |  |  | G3: sandblasting (Al_2_O_3_) | adhesive (Adper Single Bond 2) (total-etch) |  | 55.56 ± 14.76 |
| Peterson et al. (2017) | Venus Diamond | Constic  Fusio Liquid Dentin  Vertise Flow  Venus Diamond | thermal cycling (5000 cycles; 5–55˚C)/same | 16 | G1: abrasive paper + diamond bur | adhesive (OptiBond FL) (total-etch) | SBS | (Constic): 12.7 ± 2.7  (Fusio): 19.9 ± 6.4  (Vertise): 12.0 ± 3.0  (Venus): 16.4 ± 7.4 |
|  |  |  |  |  | G2: diamond bur | No treatment |  | (Constic): 10.9 ± 3.9  (Fusio): 14.7 ± 6.2  (Vertise): 6.5 ± 6.2  (Venus): 5.9 ± 3.3 |
|  |  |  |  |  | G3: abrasive paper + Sandblasting (Al_2_O_3_) | adhesive (OptiBond FL) (total-etch) |  | (Constic): 11.7 ± 3.0  (Fusio): 15.1 ± 2.7  (Vertise): 14.2 ± 3.3  (Venus): 17.3 ± 5.1 |
|  |  |  |  |  | G4: abrasive paper + Sandblasting (Al_2_O_3_) | No treatment |  | (Constic): 10.4 ± 2.1  (Fusio): 18.8 ± 5.2  (Vertise): 10.0 ± 3.3  (Venus): 16.6 ± 4.5 |
|  |  |  |  |  | G5: abrasive paper + Silica coating | adhesive (OptiBond FL) (total-etch) |  | (Constic): 10.3 ± 3.8  (Fusio): 19.8 ± 5.3  (Vertise): 14.4 ± 4.1  (Venus): 21.3 ± 3.1 |
|  |  |  |  |  | G6: abrasive paper + Silica coating | No treatment |  | (Constic): 13.5 ± 3.6  (Fusio): 21.7 ± 7.4  (Vertise): 12.2 ± 3.9  (Venus): 24.4 ± 2.8 |
|  |  |  |  |  | G7: abrasive paper | adhesive (OptiBond FL) (total-etch) |  | (Constic): 10.0 ± 4.3  (Fusio): 15.9 ± 5.5  (Vertise): 9.1 ± 2.4  (Venus): 11.0 ± 5.7 |
|  |  |  |  |  | G8: abrasive paper | No treatment |  | (Constic): 5.6 ± 3.4  (Fusio): 6.4 ± 4.1  (Vertise): 6.5 ± 4.0  (Venus): 0.3 ± 0.6 |
| Souza et al. (2017) | Esthet-X | Esthet-X | 1 year in artificial saliva/same | 16 | G1: No treatment | No treatment | µTBS | 11.18 ± 4.86 |
|  |  |  |  |  | G2: Sandblasting (Al_2_O_3_) | No treatment |  | 30.48 ± 13.86 |
|  |  |  |  |  | G3: Sandblasting (Al_2_O_3_) | adhesive (Adper Scotchbond Multi-Purpose) (total-etch) |  | 34.96 ± 20.74 |
|  |  |  |  |  | G4: Sandblasting (Al_2_O_3_) | silane |  | 30.67 ± 12.81 |
| Kiomarsi et al. (2017) (a) | Filtek Z250 | Filtek Z250 | thermal cycling (5000 cycles; 5–55˚C; dwell time: 20 s)/same | 10 | G1: diamond bur | silane | SBS | 13.85 ± 2.50 |
|  |  |  |  |  | G2: diamond bur | silane + adhesive (Adper Single Bond 2) (total-etch) |  | 20.54 ± 4.14 |
|  |  |  |  |  | G3: diamond bur | silane- + adhesive (Single Bond Universal) (total-etch) |  | 28.29 ± 4.35 |
|  |  |  |  |  | G4: Er;Cr:YSGG laser | silane |  | 4.75 ± 1.73 |
|  |  |  |  |  | G5: Er;Cr:YSGG laser | silane + adhesive (Adper Single Bond 2) (total-etch) |  | 16.05 ± 2.13 |
|  |  |  |  |  | G6: Er;Cr:YSGG laser | silane + adhesive (Single Bond Universal) (total-etch) |  | 24.61 ± 3.51 |
| Kiomarsi et al. (2017) (b) | Filtek Z250 | Filtek Z250 | thermal cycling (10000 cycles; 5–55˚C; dwell time: 20 s)/ same (5000 cycles) | 10 | G1: diamond bur | silane + adhesive (Adper Single Bond 2) (total-etch) | SBS | 19.08 ± 3.51 |
|  |  |  |  |  | G2: Er,Cr:YSGG laser | silane + adhesive (Adper Single Bond 2) (total-etch) |  | 14.08 ± 2.90 |
| Wiegand et al. (2015) | Filtek Supreme  XTE | Filtek Supreme  XTE | thermal cycling (5000 cycles; 5–55˚C; dwell time: 20 s; transfer time: 10 s)/same | 12 | G1: abrasive paper | adhesive (OptiBond FL) (total-etch) | SBS | 8.8 ± 2.7 |
|  |  |  |  |  | G2: abrasive paper + diamond bur | adhesive (OptiBond FL) (total-etch) |  | 23.8 ± 5.5 |
|  |  |  |  |  | G3: abrasive paper + sandblasting (Al_2_O_3_) | adhesive (OptiBond FL) (total-etch) |  | 19.9 ± 4.6 |
|  |  |  |  |  | G4: abrasive paper + silica coating | silane + adhesive (OptiBond FL) (total-etch) |  | 21.8 ± 6.5 |
| Musa and Nahedh (2014) | Aelite LS  Posterior | Filtek Z250 | 4 weeks in water/thermal cycling (5000 cycles; 5–55˚C; dwell time: 5 s) | 12 | G1: No treatment | No treatment | SBS | (Aelite): 13.77 ± 4.14 |
|  |  |  |  |  | G2: No treatment | adhesive (One Step Plus) (total-etch) |  | (Aelite):15.80 ± 4.63 |
|  |  |  |  |  | G3: No treatment | adhesive (Adper Single Bond 2) (total-etch) |  | (Filtek Z250): 19.61 ± 1.22 |
|  | Filtek Z250 | Filtek Z250 |  | 12 | G4: No treatment | adhesive (Adper Single Bond 2) (total-etch) |  | (Filtek Z250): 17.79 ± 2.64 |
| Eliasson et al. (2014) | Tetric Evo Ceram | Tetric Evo Ceram | thermal cycling (5000 cycles; 5–55˚C; dwell time: 20 s; transfer time: 3 s)/same thermocycling + 12 months in water | 44 | G1: No treatment | No treatment | µTBS | (cohesive): 49.6 ± 5.1 |
|  |  |  |  | 52 | G2: abrasive paper | adhesive (AdheSE One) (self-etch) |  | 24.1 ± 7.3 |
|  |  |  |  | 40 | G3: abrasive paper | adhesive (Clearfil SE Bond) (self-etch) |  | 33.6 ± 8.4 |
|  |  |  |  | 40 | G4: abrasive paper | adhesive (Adper Scotchbond Multi-Purpose) (total-etch) |  | 21.2 ± 9.9 |
|  |  |  |  | 41 | G5: abrasive paper + silica coating | adhesive (AdheSE One) (self-etch) |  | 32.9 ± 8.5 |
|  |  |  |  | 61 | G6: abrasive paper + silica coating | adhesive (Clearfil SE Bond) (self-etch) |  | 36.8 ± 10.7 |
|  |  |  |  | 58 | G7: abrasive paper + silica coating | adhesive (Adper Scotchbond Multi-Purpose) (total-etch) |  | 30.4 ± 8.3 |
|  |  |  |  | 57 | G8: abrasive paper | silane + adhesive (AdheSE One) (self-etch) |  | 33.8 ± 6.6 |
|  |  |  |  | 59 | G9: abrasive paper | silane + adhesive (Clearfil SE Bond) (self-etch) |  | 41.3 ± 7.5 |
|  |  |  |  | 53 | G10: abrasive paper | silane + adhesive (Adper Scotchbond Multi-Purpose) (total-etch) |  | 28.2 ± 6.2 |
| El-Askary et al. (2012) | Grandio Caps | Grandio Caps | 1 month in water/same | 10 | G1: diamond bur | adhesive (Solobond Plus) (total-etch) (wet bonding) | TBS | 16.9 ± 5.3 |
|  |  |  |  |  | G3: diamond bur | silane - adhesive (Solobond Plus) (total-etch) (wet bonding) |  | 12.3 ± 2.5 |
| Staxrud and Dahl (2011) | Filtek Z 250  Charisma  Filtek Supreme XT  CeramX Mono  Tetric Evo Ceram | Filtek Z 250  Charisma  Filtek Supreme XT  CeramX Mono  Tetric Evo Ceram | 60 days in water (37˚C)/thermal cycling (5000 cycles; 5–55˚C; dwell time: 20 s; transfer time: 2-3 s) | 10 | G1: abrasive paper | adhesive (Adper Scotchbond MP) (total-etch) | SBS | Filtek Z 250: 22.7 ± 4.3 |
|  |  |  |  |  | G2: abrasive paper | No treatment |  | charisma: 16.8 ± 4.7 |
|  |  |  |  |  | G3: abrasive paper | adhesive (Adper Scotchbond 1XT) (total-etch) |  | Filtek Supreme XT: 19.0 ± 5.6 |
|  |  |  |  |  | G4: abrasive paper | adhesive (Xeno III) (self-etch) |  | CeramX Mono: 16.6 ± 4.4 |
|  |  |  |  |  | G5: abrasive paper | adhesive (AdheSE) (total-etch) |  | Tetric Evo Ceram: 16.0 ± 9.8 |
| Papacchini et al. (2007) | Gradia Direct Anterior | Gradia Direct Anterior; | 1 month in 0.9% saline (37˚C)/thermal cycling (5000 cycles; 5–55˚C; dwell time: 30 s; transfer time: 5 s) | 5 | G1: sandblasting (Al_2_O_3_) | adhesive (Adper Scotchbond Multi-Purpose) (total-etch) | µTBS | 32.8 ± 6.6 |
|  |  |  |  |  | G2: sandblasting (Al_2_O_3_) | flowable resin composites (Filtek Supreme XT Flow) |  | 48.2 ± 7.5 |
|  |  |  |  |  | G3: sandblasting (Al_2_O_3_) | flowable resin composites (Gradia LoFlo) |  | 43.9 ± 7.7 |
|  |  |  |  |  | G4: sandblasting (Al_2_O_3_) | silane |  | 28.3 ± 9.6 |
|  |  |  |  |  | G5: sandblasting (Al_2_O_3_) | silane + adhesive (Clearfil New Bond) (total-etch) |  | 34.7 ± 11.2 |
|  |  |  |  |  | G6: sandblasting (Al_2_O_3_) | silane + adhesive (Clearfil SE Bond) (self-etch) |  | 38.0 ± 9.5 |
|  |  |  |  |  | G7: sandblasting (Al_2_O_3_) | silane + adhesive (Clearfil Tri-S Bond) (self-etch) |  | 33.7 ± 13.0 |
