## Supplementary figures and images for "Which surface treatment improves the long-term repair bond strength of aged methacrylate-based composite resin restorations? A systematic review and network meta-analysis"

### Supplementary material 5. Risk of bias assessment of individual studies.

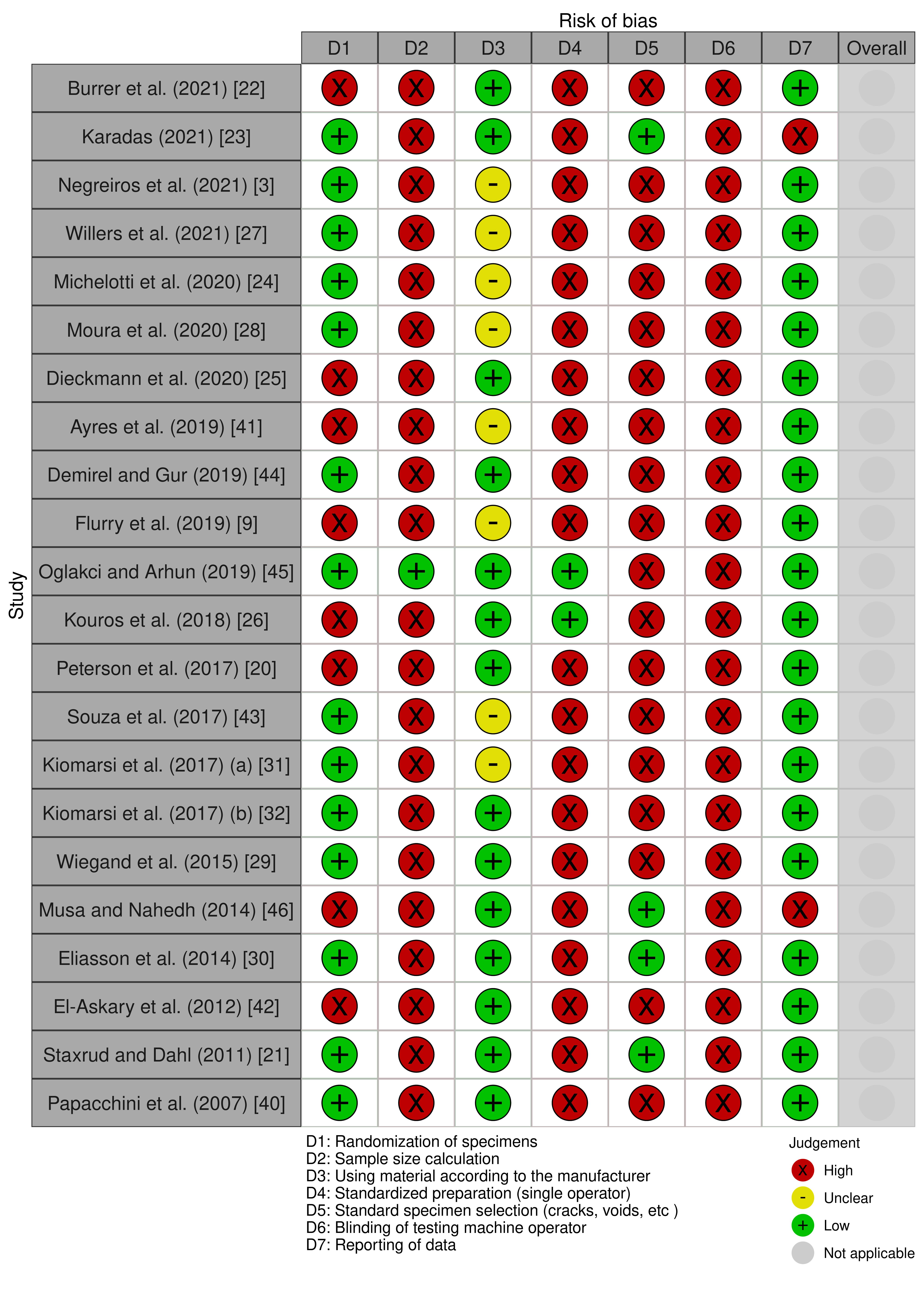

### Supplementary material 6. League tables (two-by-two comparisons).

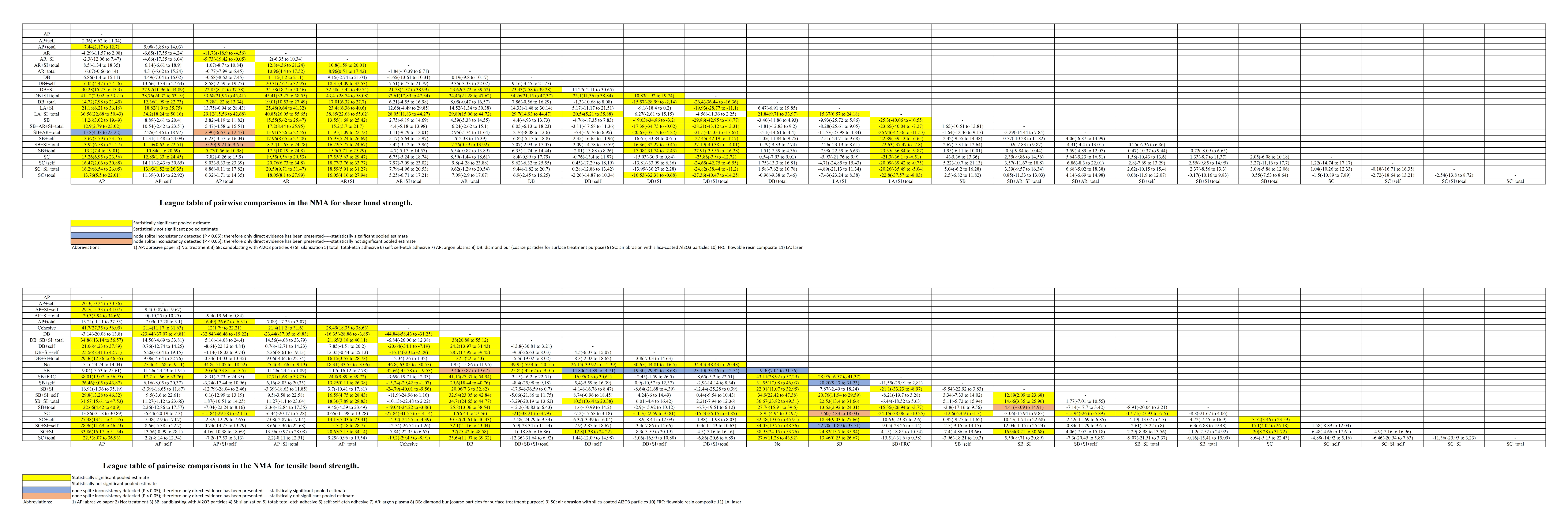

### Supplementary material 7. Publication bias (funnel plots).

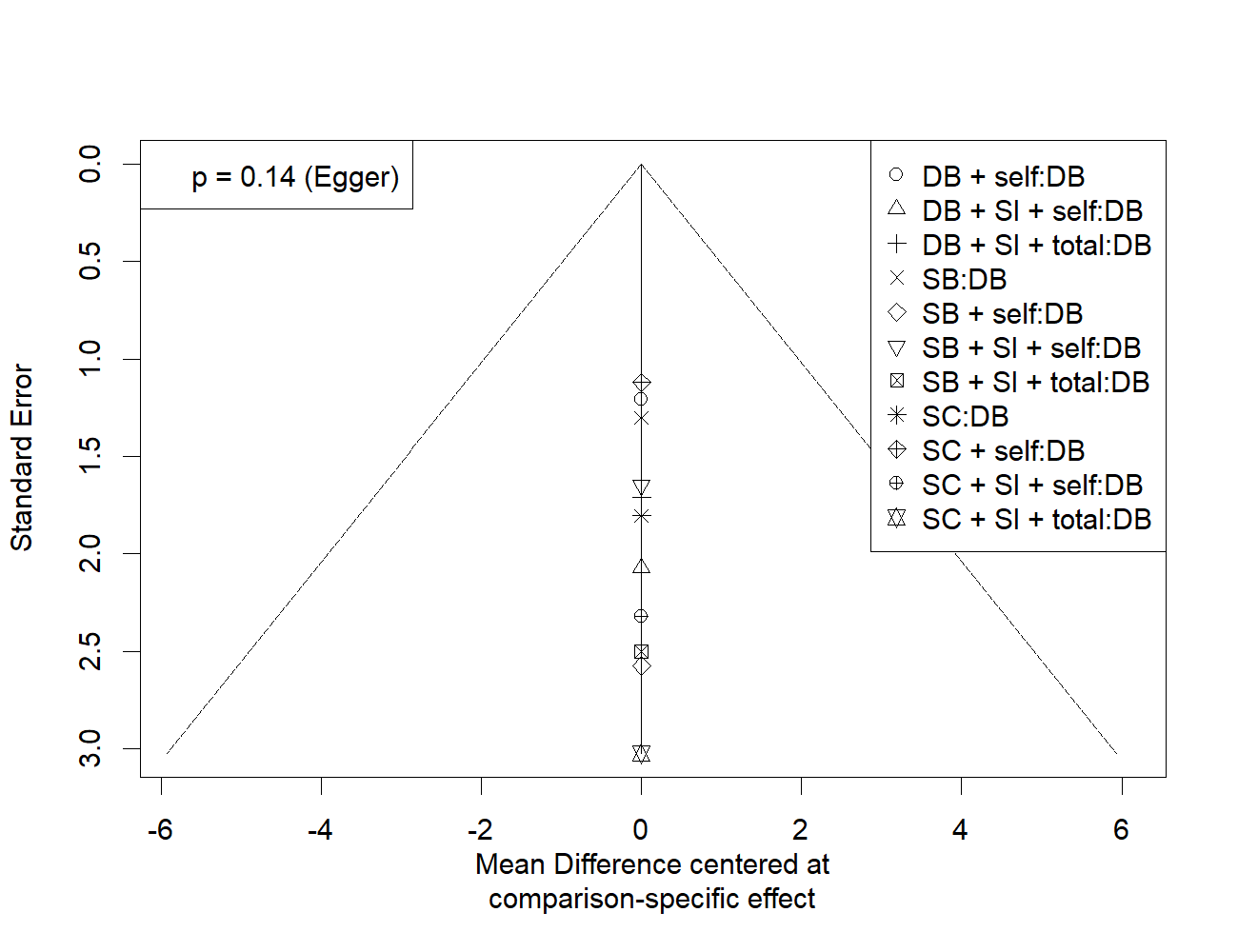

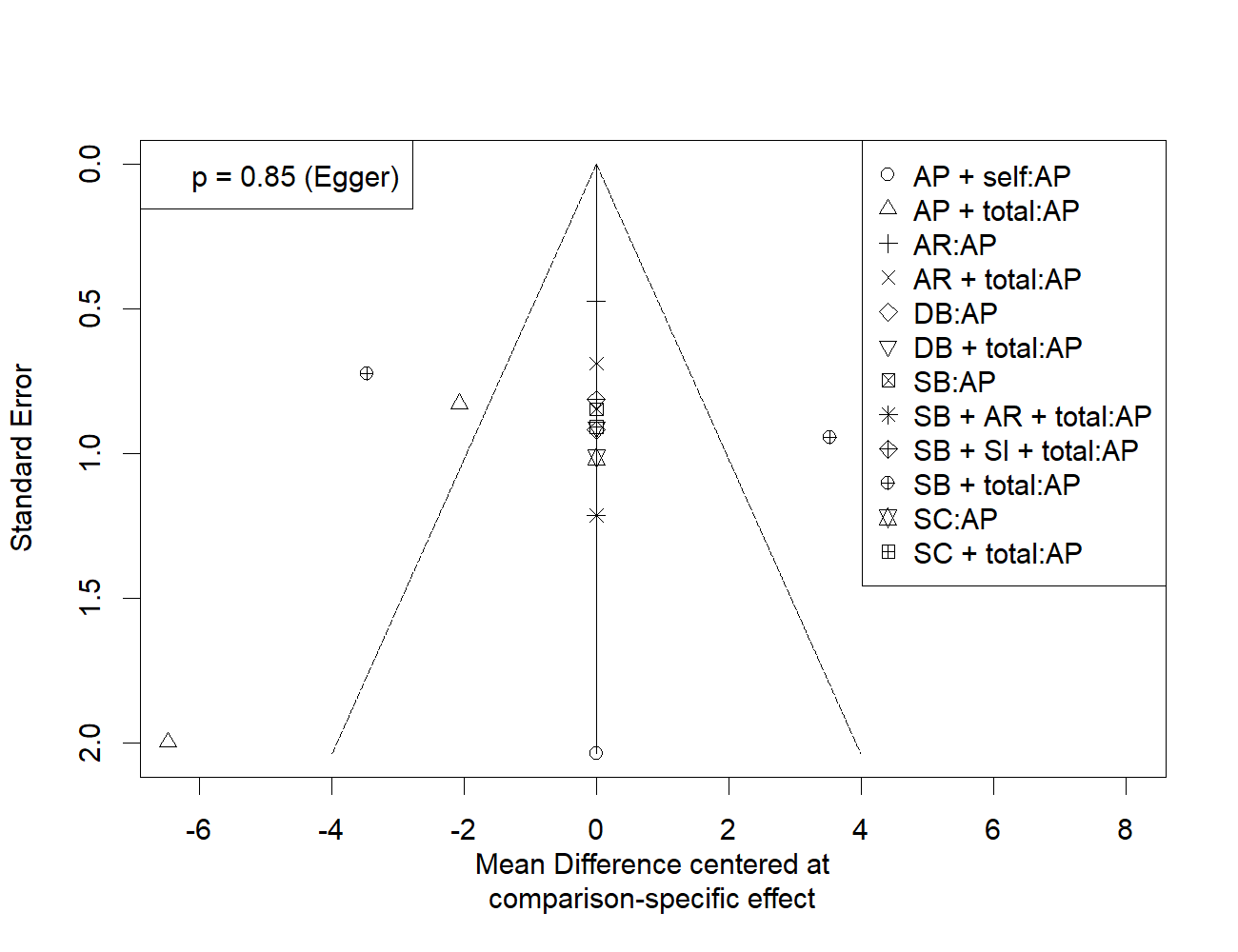

### Supplementary material 8. Node-split inconsistency results.

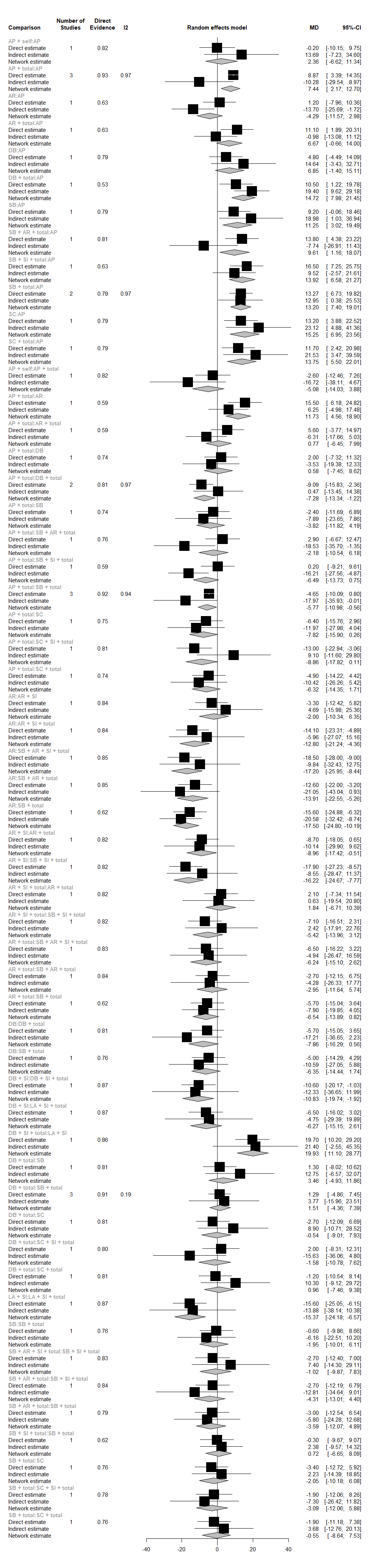

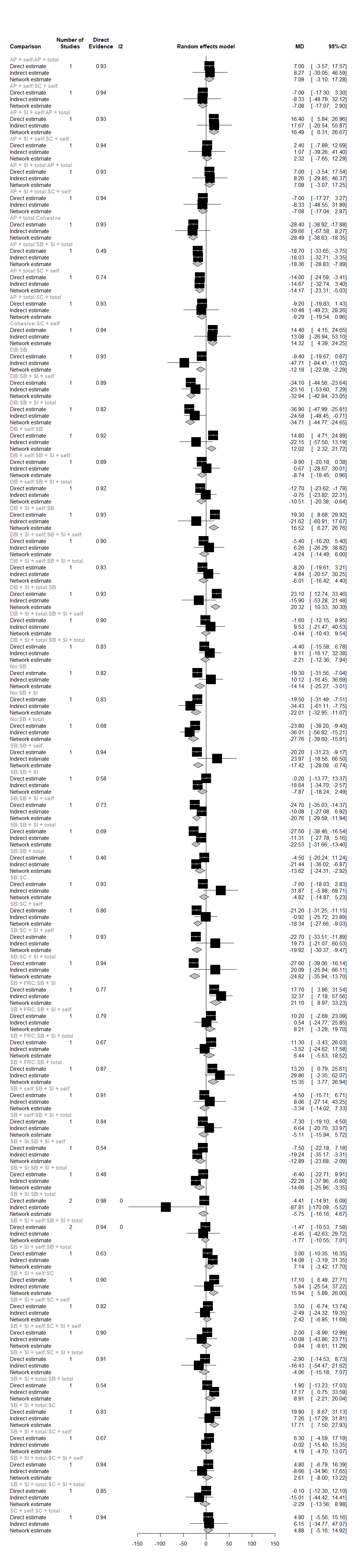
sssss

(micro)Shear

(micro)Tensile
